# The role of sleep in the human brain and body: insights from multi-organ imaging genetics

**DOI:** 10.1101/2022.09.08.22279719

**Authors:** Zirui Fan, Yilin Yang, Yuxin Guo, Yujue Li, Juan Shu, Xiaochen Yang, Bingxuan Li, Jinjie Lin, Qianwen Wang, Carolyn Gibson, Peristera Paschou, Tengfei Li, Philip Gehrman, Hongtu Zhu, Bingxin Zhao

## Abstract

Sleep is crucial for overall physical and mental health, concerning organs such as the brain, heart, eye, liver, kidney, and lung. Nonetheless, a thorough understanding of how sleep relates to these organs, as well as their genetic bases, remains elusive. Here we conducted a systematic mapping of sleep-organ connections, focusing on 623 multi-organ imaging biomarkers and 10 sleep traits. Both phenotypic and genetic analyses uncovered robust associations between sleep and the structure and function of multiple organs, such as brain functions measured by functional magnetic resonance imaging (fMRI) and body composition traits in abdominal MRI. Sleep and imaging traits had shared genetic influences in 51 genomic loci, 23 of which had colocalized causal genetic effects. Considering the extensive genetic overlaps between sleep and multi-organ imaging biomarkers, we further described the patterns of genetic similarities between sleep and 50 diseases relevant to these organs. Notably, among all diseases examined, psychiatric disorders consistently exhibited the strongest genetic correlations and causal links with sleep. Mediation analysis further revealed that many of the identified sleep-imaging connections were mediated by intra or inter-organ diseases. Overall, our study demonstrates the broad implications of sleep on brain and body health, influenced in part by shared genetic factors.

## Background

Sleep has a pivotal role in physical and mental health^1^. Sleep traits (such as sleep duration) and disorders (such as insomnia and narcolepsy) are associated with a wide spectrum of neurodegenerative diseases, psychiatric disorders, cardiovascular diseases, metabolic syndromes, as well as diseases related to the liver, lung, and kidney. For example, sleep dysregulation and short sleep duration in midlife have been linked to a higher risk of Alzheimer’s disease and other dementias^2^. Individuals with sleep disturbance were also more likely to have mental and psychiatric disorders^3^, such as anxiety^4^ and schizophrenia^5^. In addition, both insufficient and excess sleep duration may cause a higher incidence of cardiovascular outcomes, including coronary heart disease^6^, atrial fibrillation^7^, and stroke^8^. Sleep and circadian rhythm disorders, including obstructive sleep apnea (OSA) and circadian rhythm, have an impact on metabolic syndromes such as obesity and type 2 diabetes^9–11^. Moreover, OSA leads to insulin resistance and dyslipidemia, which play an important role in the development of nonalcoholic fatty liver disease (NAFLD)^12,13^. Sleep complaints, including insomnia and OSA, were frequently encountered in patients with chronic kidney disease^14,15^, as well as lung diseases such as chronic obstructive pulmonary disease (COPD)^16,17^ and asthma^16^. However, achieving a thorough understanding of sleep from a pan-organ perspective remains a challenge. Two methodological approaches that can be used to investigate the relationships between sleep and organ function are imaging and genetics.

Magnetic resonance imaging (MRI) provides a noninvasive and comprehensive measure of organ structure and function. Many imaging traits extracted from brain, cardiac, and abdominal MRI are well-established clinical endophenotypes and have been widely used in the prediction and early detection of complex diseases^18–24^. In addition, retinal optical coherence tomograph (OCT) imaging is a non-invasive approach that offers a high-resolution view of the cross-sectional structure of the retina^25^. Using MRI and OCT data, several studies have examined the sleep-related changes in structures and functions of the brain^26–31^, heart^32,33^, and eye^34–36^. For example, poor sleep quality is linked to reduced hippocampal volume^26^ and brain atrophy in the right superior frontal cortex^27^, as well as decreased functional connectivity in several networks, including those in the left middle temporal and left inferior occipital regions^29^. Cardiac MRI suggests that individuals with OSA tend to exhibit lower ventricular volumes and increased ejection fractions^33^.

Additionally, OSA is associated with thicker choroidal thickness and thinner mean RNFL thickness in both eyes, as observed by OCT^34^. Three major limitations of most existing studies have been i) the limited study sample size, which was usually less than a few hundred; ii) focusing on one organ, single imaging modality (or trait), and/or one sleep trait, such as hippocampal atrophy^26^ and sleep duration^31,37,38^; and iii) a scarcity of investigations on sleep-related changes in abdominal organs using MRI. It is known, however, that large sample sizes are needed for imaging studies to detect small effect sizes and produce reproducible findings^39^. In addition, distinct imaging modalities and sleep traits may be relevant to different diseases and health-related characteristics^40^. Therefore, a systematic analysis of multi-organ imaging data and multiple sleep traits would provide a more extensive map of sleep-related structural and functional variations across multiple organs.

Large-scale genome-wide association studies (GWAS) have shown that sleep disorders and traits are heritable and have a polygenic genetic architecture^41–56^. The sleep-associated genetic variants are enriched for genes expressed in the brain and for metabolic and psychiatric pathways^55^. Genetic correlations between sleep traits and brain-related disorders (such as depression and schizophrenia) have been discovered, suggesting their shared neurogenetic basis^57^. On the other hand, imaging traits are also heritable and hundreds of associated genetic loci have been identified in recent GWAS^58–70^. However, few studies have ever integrated these heritable multi-organ health indexes and sleep traits to explore the genetic interactions among sleep, organ structure/function, and associated clinical endpoints.

To better understand sleep’s role in the brain and overall body health, we examined the phenotypic and genetic connections between sleep and multi-organ imaging traits of the brain, heart, abdomen (such as liver, kidney, and lung), and retina obtained from the UK Biobank (UKB) study^71^. We used 623 traits derived from various imaging modalities, including brain structural MRI^65^, brain diffusion MRI^67^, brain resting and task functional MRI^69^, cardiac MRI^72^, abdominal MRI^22–24,73–79^, and retinal OCT^70^ (**Table S1**). We examined 7 self-reported sleep-related phenotypes (sleep duration, getting up in the morning, chronotype, sleeplessness/insomnia, nap during the day, narcolepsy, and snoring), along with 3 accelerometer-derived measurements of sleep duration. We further examined the genetic connections between sleep and 50 relevant diseases of the organs studied, with 113 sets of their publicly available GWAS summary statistics. The overview of our study is presented in **Figure 1**.

**Fig. 1.**
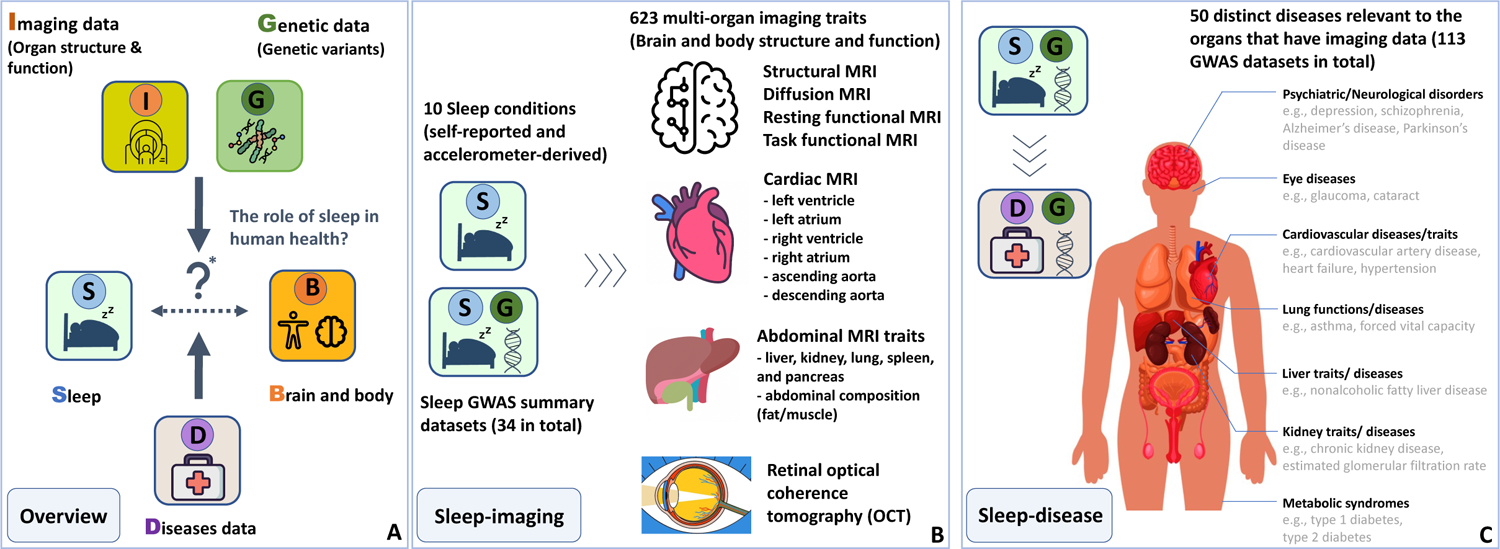
Study overview. **(A)** Our study aimed to demonstrate the connections between sleep traits and brain and body health using multi-organ imaging, genetic, and disease data. We assessed organ structure and function through imaging traits and considered their related clinical outcomes. **(B)** Encompassing a diverse range of imaging modalities, our study included brain imaging traits such as structural MRI, diffusion MRI, resting and task fMRI; cardiac MRI such as short-axis, long-axis, and aortic cine images; abdominal MRI such as derived measurements of liver, kidney, lung, pancreas, spleen, and abdominal fat/muscle composition; and retinal derived OCT measures. Multiple sleep traits were examined, including ease of getting up in the morning, chronotype, nap during the day, insomnia, snoring, narcolepsy, as well as self-reported and accelerometer-derived sleep duration measures. We used individual-level imaging and sleep data from the UKB study, as well as the publicly available sleep GWAS summary statistics. **(C)** In addition, we examined 50 clinical outcomes related to the organs that have imaging data.

## RESULTS

### Sleep-imaging atlas: phenotypic and genetic connection patterns across multiple organs

We examined the phenotypic associations between 10 sleep traits (7 self-reported and 3 accelerometer-derived) and a wide range of imaging traits across multiple organs, including the brain, heart, eye, liver, lung, kidney, pancreas, and spleen, as well as body muscle/fat composition. The discovery analysis used data from unrelated white British subjects, and we reported significant associations at a 5% false discovery rate (FDR) using the Benjamini-Hochberg procedure (*P* range = (7.17×10^-87^, 1.60×10^-2^), **Table S2**). To examine the robustness and reproducibility of these findings, two approaches were used: adjusting for a wide range of additional covariates based on literature and replicating the analysis in an independent hold-out dataset (**Methods**). Below, we present associations in the discovery sample that remained nominal significance level (*P* < 0.05) in both approaches.

Among all brain MRI modalities, resting fMRI measures of brain function exhibited the strongest phenotypic associations with sleep traits (**Figs. 2A** and **S1**). Resting fMRI traits were associated with various sleep traits, particularly with sleep duration, narcolepsy, and ease of getting up in the morning (**Fig. S2**). Both self-reported and accelerometer-derived sleep duration traits had consistent associations with resting fMRI traits (β range = (−0.17, 0.10), *P* < 4.43×10^-4^). Most of these associations were negative and predominantly linked to the somatomotor and visual networks. The importance of somatomotor function in various sleep stages is well-established, and there is a consistent correlation between poor sleep quality, shorter sleep duration, and increased functional connectivity in the somatomotor network among young individuals^80–82^. In addition, increased resting functional connectivity was primarily associated with an elevated risk of narcolepsy (β range = (−0.04, 0.07), *P* < 9.24×10^-4^) and a greater ease of getting up in the morning (β range = (−0.02, 0.05), *P* < 6.23×10^-3^). The resting fMRI traits linked to narcolepsy and the ease of getting up were largely a subset of those associated with sleep duration, with the somatomotor and visual networks again being emphasized, highlighting the crucial role these two networks play in sleep regulation. In contrast to resting fMRI, task fMRI overall had weaker connections with sleep, with the most significant associations being observed with accelerometer-derived sleep duration (β range = (−0.08, −0.02), *P* < 1.60×10^-2^, **Fig. S3**). Almost all significant associations with sleep duration traits were negative and showed a substantial overlap with those identified in resting fMRI.

**Fig. 2.**
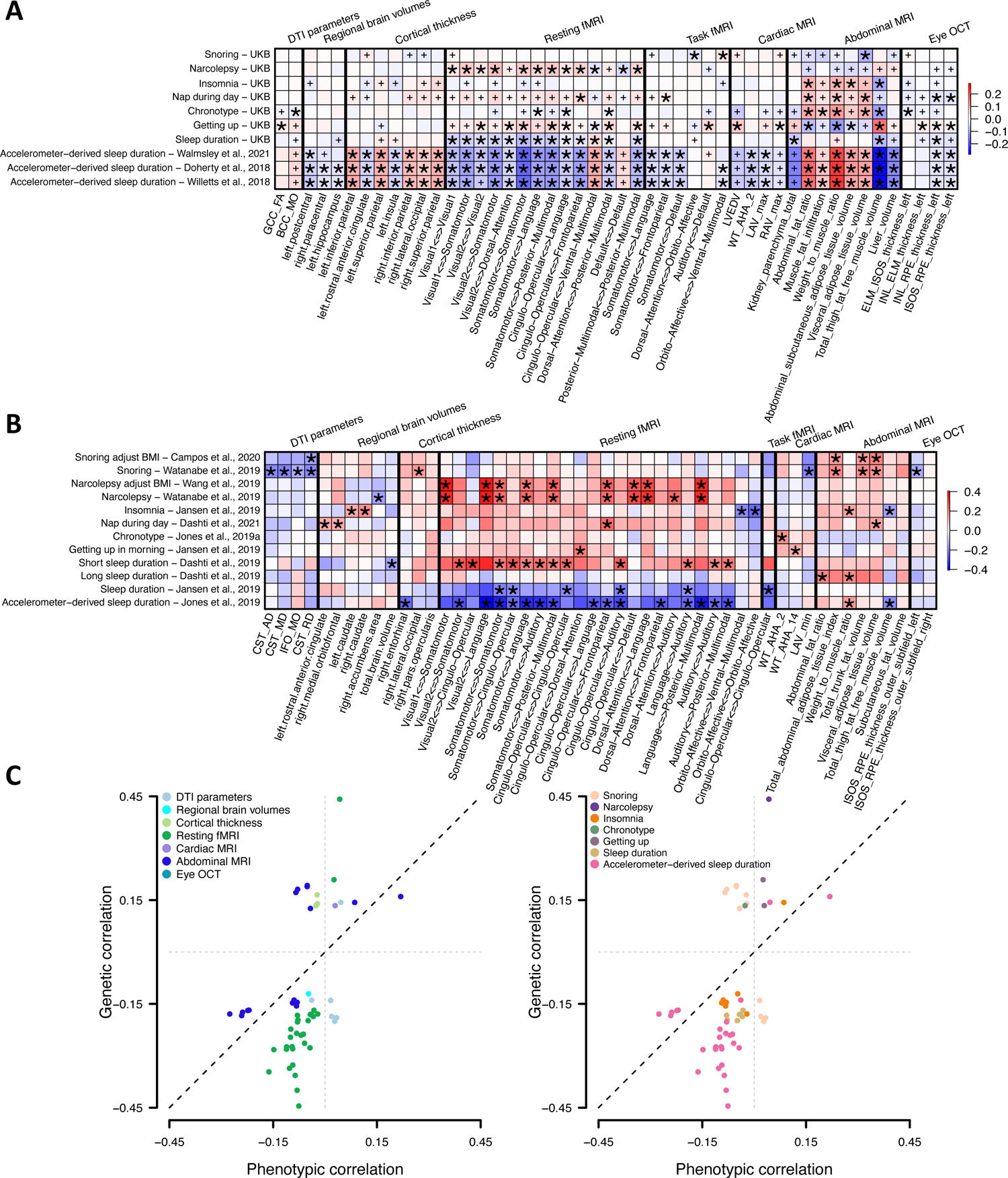
Phenotypic and genetic sleep-imaging associations. **(A)** Selected phenotypic associations between sleep traits and multi-organ imaging traits. The color represents correlation estimates. Associations that passed the FDR rate of 5% (*P* < 1.60×10^-2^) in the discovery sample and survived the two validation approaches were marked with an asterisk. Associations that passed the FDR rate of 5% in the discovery sample but failed in any of the two validation approaches were marked with a plus sign. See **Table S1** for more information on imaging traits. **(B)** Selected genetic correlations between sleep and multi-organ imaging traits. The color represents correlation estimates. Associations that passed the FDR rate of 5% (*P* < 5.08×10^-4^) were marked with an asterisk. **(C)** Sleep-imaging pairs were significant in both phenotypic and genetic correlation analyses at a 5% FDR level. We illustrate genetic correlation (*y*-axis) and phenotypic correlation (*x*-axis) between sleep and imaging traits. The color represents imaging categories (left panel) or sleep traits (right panel).

Associations detected in brain structural MRI traits, such as cortical thickness and regional brain volumes, were mostly correlated with accelerometer-derived sleep duration (**Figs. S4** and **S5**). For example, sleep duration was associated with the thickness of left/right rostral anterior cingulate, inferior parietal, and insula (β range = (−0.07, 0.10), *P* < 4.86×10^-3^), as well as the volume of left paracentral, postcentral, and hippocampus (β range = (− 0.05, 0.04), *P* < 1.09×10^-2^). For diffusion tensor imaging (DTI) parameters derived from brain diffusion MRI, differences in white matter of the body of corpus callosum and genu of corpus callosum tracts were linked to the ease of getting up in the morning (β range = (0.021, 0.023), *P* < 6.35×10^-4^, **Fig. S6**), which was consistent with previous studies that being an evening person exhibited differences in several white matter tracts, including the corpus callosum^83,84^.

Imaging traits from other organs also had strong connections with sleep, especially for abdominal MRI traits. Abdominal MRI traits were associated with multiple sleep traits, including sleep duration, insomnia, daytime napping, and chronotype (**Fig. S7**). Longer sleep duration was linked to reduced renal parenchymal volume and liver volume (β range = (−0.13, −0.04), *P* < 1.45×10^-7^), as well as multiple body and abdominal fat traits, such as anterior/posterior thigh fat-free muscle volume (FFMV), anterior/posterior thigh muscle fat infiltration, visceral adipose tissue volume (VAT), and subcutaneous fat volume (β range = (−0.28, 0.22), *P* < 2.63×10^-6^). Many of these body/abdominal fat measurements were also associated with other sleep traits. For example, an increase in abdominal fat ratio, total abdominal adipose tissue index, weight to muscle ratio, and total trunk fat volume was associated with elevated risk of insomnia, nap during the day, evening chronotype (β range = (0.06, 0.11), *P* < 1.47×10^-11^), and difficulty getting up in the morning (β range = (−0.11, −0.05), *P* < 3.82×10^-8^). Disrupted sleep patterns, particularly short/long sleep durations, can lead to inflammation and metabolic disorders and further contribute to obesity and an increased risk of liver cancer or chronic liver disease^85–87^. The physiological processes of the kidneys were influenced by the circadian rhythms, and increasing evidence indicated that abnormal sleep durations affect reduced renal function and the development of kidney-related diseases^88–90^. Our findings shed light on the potential mechanisms through which sleep duration impacts the onset of those diseases related to the liver and kidney.

Furthermore, retinal OCT traits were associated with many sleep traits (**Fig. S8**). For example, reduced average thickness between the inner nuclear layer (INL) to the retinal pigment epithelium (RPE), as well as between the inner and outer photoreceptor segments (ISOS) to RPE were associated with longer accelerometer-based and self-reported sleep duration, napping during the day, and difficulty in getting up in the morning (β range = (−0.04, 0.03), *P* < 6.43×10^-3^). Many previous studies have examined the relationship between retinal thickness and OSA^34–36^, but there are relatively few studies focused on other sleep traits. For cardiac MRI, we observed links between sleep duration and myocardial-wall thickness at end-diastole, left atrium maximum volume, and left atrium stroke volume (**Fig. S9**, β range = (−0.07, 0.03), *P* < 6.40×10^-3^), which was consistent with previous findings that sleep duration is a potential factor for cardiac remodeling^91,92^.

To evaluate the sleep-imaging genetic correlations, we collected 34 sets of sleep GWAS summary statistics from various studies (**Table S3**) and performed linkage disequilibrium (LD) score regression^93^ with our imaging traits. At a 5% FDR level (*P* range = (2.23×10^-14^, 5.08×10^-4^), we observed similar patterns of genetic correlation to those observed in the phenotypic association analyses, with most of the significant genetic correlations concentrated in resting fMRI and abdominal MRI traits (**Figs. 2B**, **S10-S11,** and **Table S4**). Resting fMRI had the most pronounced genetic correlation with sleep, especially for sleep duration and narcolepsy. Increased resting functional connectivity in multiple networks, such as the somatomotor, cingulo-opercular, and language, were genetically correlated with shorter sleep duration and increased risk of narcolepsy (β range = (0.17, 0.44), *P* < 5.00×10^-4^). The genetic correlations with narcolepsy still existed after adjusting for body mass index (BMI) (**Fig. 2B**). Accelerometer-based sleep duration had more genetic correlations with resting fMRI than self-reported sleep duration traits. Compared with resting fMRI, task fMRI had much weaker genetic correlations.

Genetic correlations were also observed for other brain imaging modalities. For example, snoring was genetically correlated with the corticospinal tract and the inferior fronto-occipital fasciculus (β range = (−0.20, −0.13), *P* < 3.62×10^-4^). The corticospinal tract is a major neuronal pathway carrying movement-related information from the cerebral cortex to the spinal cord. Impaired corticospinal tract was observed in poor cognitive performance and patients with obstructive sleep spnea^94^. Interestingly, after adjusting for BMI, only one of these genetic correlations of snoring remained (**Fig. 2B**), indicating that the majority of genetic associations between snoring and brain white matter might be mediated by BMI. For regional brain volumes, insomnia was genetically correlated with increased left/right caudate volume (β range = (0.14, 0.17), *P <* 9.20×10^-5^) as well as decreased total brain volume (β range = (−0.132, −0.126), *P <* 6.00×10^-5^). The caudate is a well-known functional region for sleep and modulates multiple sleep stages^95^. We also observed the negative genetic correlation between the volume of the right accumbens and narcolepsy (β = −0.14, *P* = 1.00×10^-4^). The adenosine *A*_2*A*_ receptors and dopamine *D*_2_ receptors in the accumbens area regulate the release of specific neurotransmitters that suppress arousal and wakefulness, thereby promoting sleep^96–98^.

Similar to phenotypic association analysis, abdominal imaging traits were genetically correlated with multiple sleep traits, including insomnia, sleep duration, and snoring. Elevated VAT and total abdominal adipose tissue index were genetically correlated with snoring (β range = (0.16, 0.19), *P* < 2.16×10^-4^). Different from the genetic correlations with DTI parameters, all genetic correlations of snoring with abdominal imaging traits continued to be significant after adjusting for BMI (**Fig. S11**). Insomnia and sleep duration primarily showed genetic correlations with abdominal body muscle and fat. For example, increased total and anterior/posterior thigh FFMV were genetically correlated with decreased risk of insomnia and shorter accelerometer-derived and self-reported sleep duration (β range = (−0.29, −0.13), *P* < 4.72×10^-4^). Previous studies have found that OSA has a strong association with metabolic syndromes, such as excessive visceral fat and insulin resistance^99,100^. Accumulation of visceral fat was reported to be a key risk factor for OSA among obese patients^101^. The genetic correlation between snoring and pancreas fat fraction (β = 0.19, *P* = 2.00×10^-4^) was also observed. OSA has been hypothesized as a potential risk factor for nonalcoholic fatty pancreatic disease, as the pancreas might be a target for fat deposition due to hormone imbalances caused by intermittent hypoxia, which is commonly experienced by OSA patients^102^.

In retinal OCT traits, we found a genetic correlation between snoring and a reduction in average thickness between the ISOS and RPE of the outer subfield (β range = (−0.13, − 0.12), *P* < 5.00×10^-4^). Intermittent hypoxia may alter the structure of retinal and choroidal blood vessels, potentially influencing the retinal and choroidal thickness^36^. Additionally, cardiac MRI traits showing significant genetic correlations with sleep largely related to myocardial-wall thickness and left atrium volumes, overlapping with those phenotypically linked to sleep. For example, increased regional myocardial-wall thickness at end-diastole was associated with being an evening person (β range = (0.13, 0.19), *P* < 5.00×10^-4^). Furthermore, snoring showed genetic correlations with decreased left atrium maximum/minimum volume (β range = (−0.19, −0.16), *P* < 5.00×10^-4^).

A total of 57 sleep-imaging pairs were found to be significant in both phenotypic and genetic analyses (*P* < 3.21×10^-3^, **Fig. 2C** and **Table S4**), with 45 out of these 57 pairs having the same direction of effect sizes in both analyses. Within these 45 pairs, 19 correlations were identified between sleep duration and resting fMRI traits. Moreover, both phenotypic and genetic correlations between insomnia and sleep duration with multiple abdominal MRI traits were observed, constituting 12 of the 45 pairs. For example, total thigh FFMV and weight-to-muscle ratio had genetic and phenotypic associations with both insomnia and accelerometer-derived sleep duration. Additionally, accelerometer-derived sleep duration demonstrated correlations with VAT. Interestingly, we found weaker phenotypic correlations compared to their genetic counterparts (*P* < 1.35×10^-14^). Specifically, of the 45 pairs, 39 had smaller absolute phenotypic correlation estimates compared to the absolute values of their corresponding genetic correlation estimates, with a few exceptions being observed between accelerometer-derived sleep duration and abdominal MRI traits (such as thigh FFMV, weight-to-muscle ratio, and abdominal fat ratio). Overall, these results indicate the substantial shared genetic influences on the sleep-imaging connections. Considering the polygenic nature of sleep and imaging traits, we aim to identify the specific genomic regions contributing to their genetic similarities in the following section.

### Characterizing sleep-imaging genetic overlaps in 51 genomic loci

To understand the genetic co-architecture underlying sleep-imaging associations, we characterized the genetic pleiotropy between sleep and multi-organ MRI traits using the NHGRI-EBI GWAS catalog^103^ (**Methods**). We found that a total of 51 independent genomic loci showed shared genetic influences on both sleep and imaging traits across multiple organs (LD *r*^2^ ≥ 0.6). Imaging-associated genetic variants were also reported in previous GWAS for a wide range of sleep traits, including insomnia^45^, chronotype^104^, daytime nap^45^, sleep problems (depressive symptom)^105^, getting up^45^, hypersomnia^106^, sleep duration^43^, and snoring^56^ (**Fig. 3** and **Table S5**). Furthermore, we estimated the probability that the sleep-imaging pairs shared causal genetic variants using the Bayesian colocalization test^107^. We considered pairs with a probability of shared causal variant (PPH4) > 0.8 to be colocalized^107,108^. Among all the pairs we tested, 23 out of the 51 regions showed evidence of shared causal variants. A complete list of these 51 regions can be found in **Table S6** and below we highlighted a few examples for each imaging modality.

**Fig. 3.**
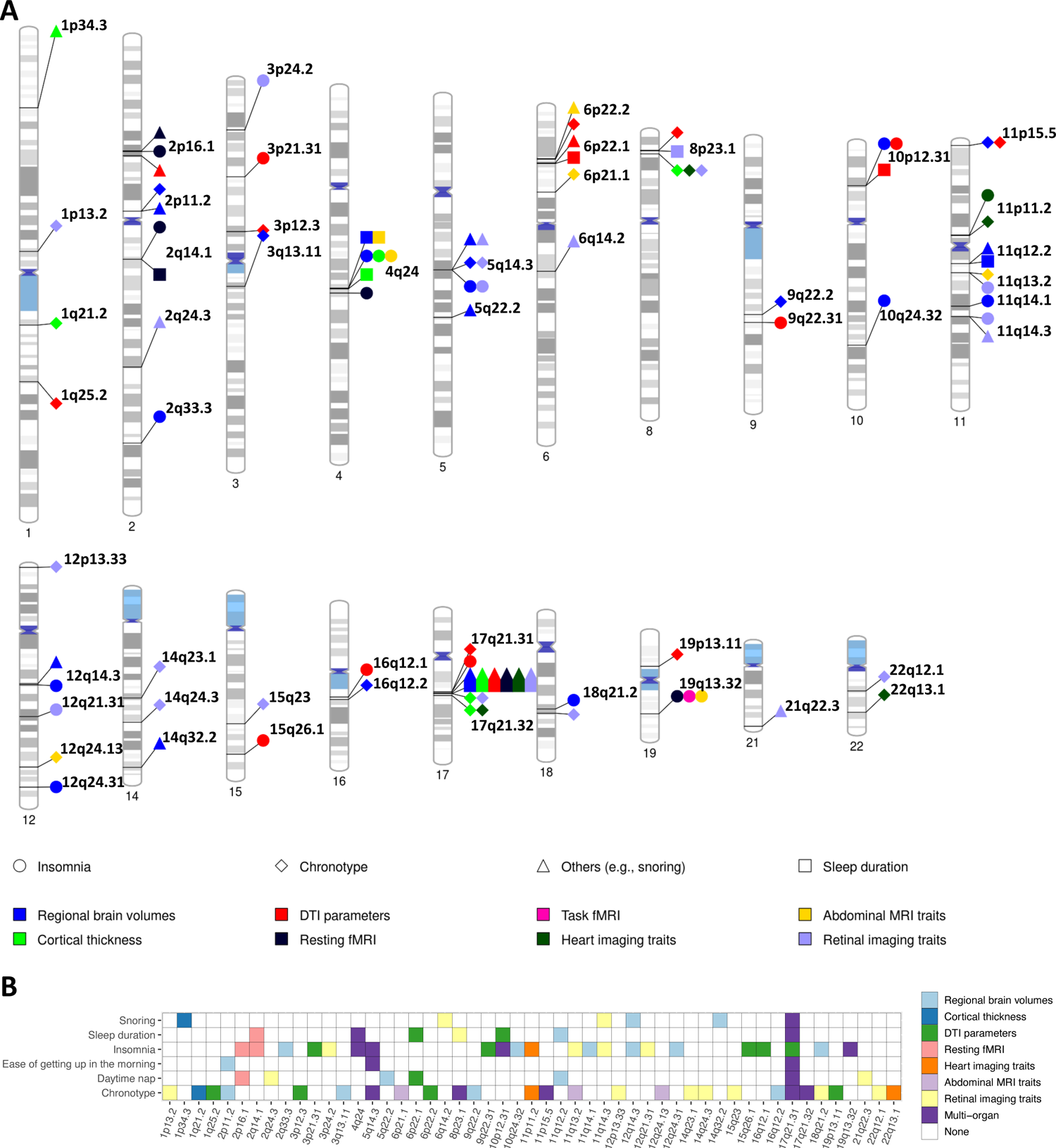
Genomic loci associated with both sleep and imaging traits. **(A)** In the NHGRI-EBI GWAS catalog, we found shared genetic influences between sleep and imaging traits in 51 genomic loci (names are in black). That is, sleep and imaging GWAS reported associated variants in these loci and their index variants were in linkage disequilibrium (LD, *r*^2^ ≥ 0.6). Each imaging modality is labeled with a different color. We tagged a wide range of reported sleep traits and grouped them into chronotype (e.g., morningness and morning person), insomnia (e.g., insomnia and insomnia symptoms), sleep duration (e.g., sleep duration long sleep and short sleep), and others, such as snoring, hypersomnia, getting up, and daytime nap. Each sleep trait is labeled with a different shape. **(B)** We further summarized the results into a table, where the *x*-axis represents the 51 genomic regions, and the *y*-axis displays the sleep traits. Each imaging modality is labeled with a different color and the dark purple color is used when more than one imaging modalities are observed in the locus.

Within different brain MRI modalities, DTI parameters and regional brain volumes exhibited numerous shared genetic loci with sleep, especially with chronotype and insomnia. There were 13 genomic loci where sleep-significant variants were associated with DTI parameters (LD *r*^2^ ≥ 0.6). For example, the corpus callosum tract had shared genetic effects with chronotype in the locus 1q25.2 (sleep index variant rs975025, **Fig. 4A**) and with insomnia in the locus 9q22.31 (rs10761240, **Fig. S12**), which was consistent with the involvement of corpus callosum in sleep regulation^83,84^. These shared loci also partially explained the associations observed between sleep and corpus callosum in our phenotypic analysis. Moreover, the internal capsule had shared genetic influences with chronotype in 6p22.2 (rs766406, **Fig. S13**), and with insomnia in 10p12.31 (rs12251016) and 15q26.1 (rs176647, **Fig. S14**). For regional brain volumes, 18 genetic regions with shared genetic influences on sleep were identified, of which three overlapped with DTI parameters. Similar to DTI parameters, the majority of these shared genetic influences were again linked to chronotype and insomnia. For example, in 2p11.2, the volume of the brain stem had shared genetic influence with ease of getting up (rs1606803, **Fig. S15**) and chronotype (rs11681299, **Fig. S16**). The variant rs11681299 is a brain expression quantitative trait locus (eQTL) of *EIF2AK3*, which encodes PERK. PERK plays a critical role in intracellular proteostatic mechanisms, essential for keeping normal sleep. It affects sleep and wake behavior by impacting the expression of wake-promoting neuropeptide^109^. Furthermore, PERK is a risk factor for neurodegenerative diseases, such as Alzheimer’s disease, due to its contribution to the pathologic aggregation of misfolded tau proteins in the nervous system^110^. Moreover, loci 11q14.1 and 18q21.2 harbor genes that influenced putamen volume, including *DLG2* in 11q14.1 and *DCC* in 18q21.2. The shared genetic effects were identified between putamen volume and insomnia in 11q14.1 (rs667730, **Fig. S17**) and chronotype in 18q21.2 (rs10502966, **Fig. S18**). These genes are highly pleiotropic for brain disorders, where *DCC* has shown associations with depression and schizophrenia in animal studies^111,112^, and *DLG2* encodes an evolutionarily conserved scaffolding protein and has been associated with schizophrenia, Parkinson’s disease, and cognitive impairment^113^. Other sleep traits also show shared genetic influences with regional brain volumes, such as in the locus 14q32.2 for snoring and volume of the left thalamus proper (**Fig. 4B**).

**Fig. 4.**
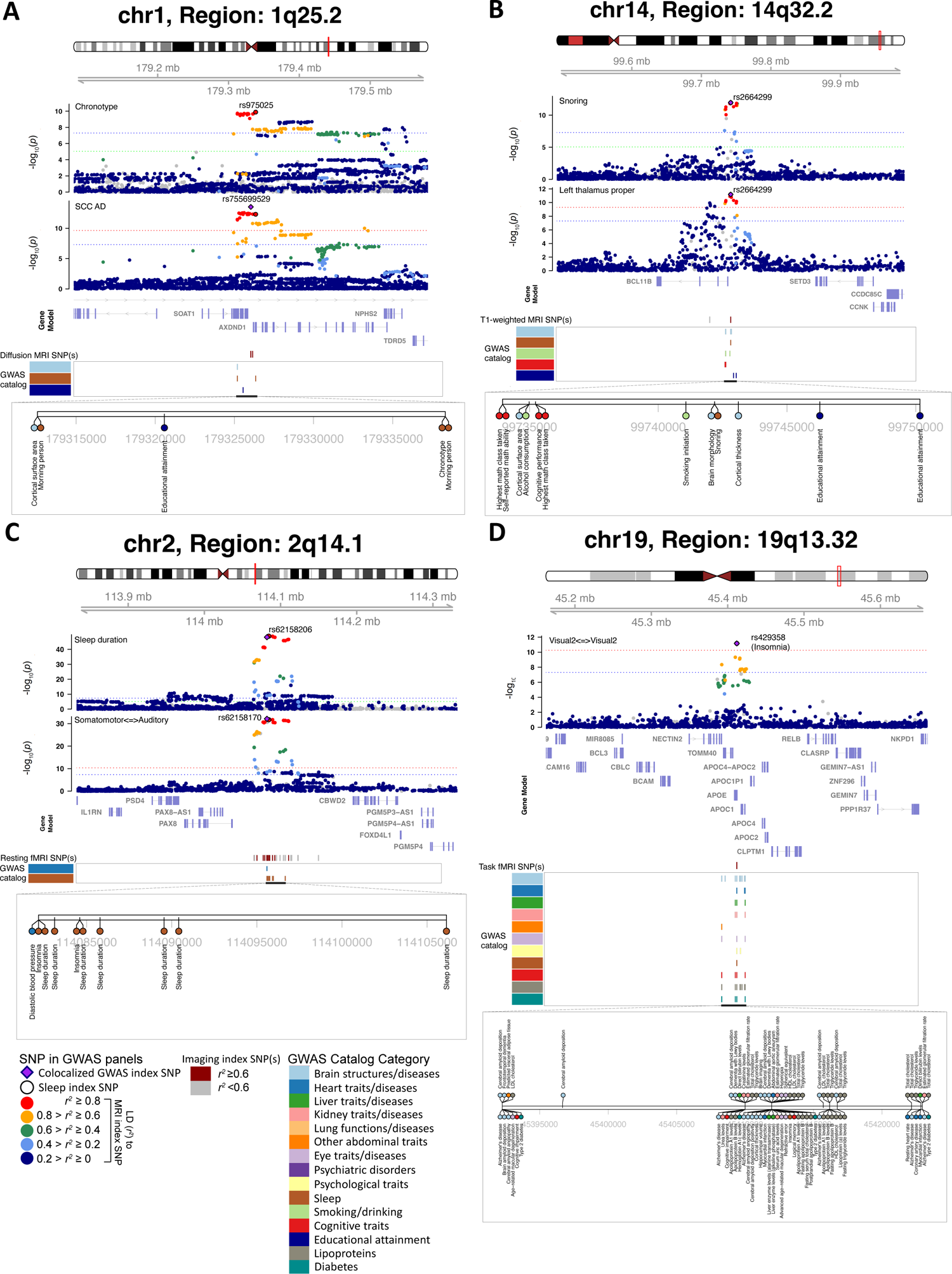
Selected genetic loci that were associated with both sleep and brain imaging traits. **(A)** In 1q25.2, we observed colocalization between chronotype (index variant rs975025) and the mean axial diffusivity of the splenium of the corpus callosum tract (SCC AD, index variant rs755699529). The posterior probability of Bayesian colocalization analysis for the shared causal variant hypothesis (PPH4) is 0.976. **(B)** In 14q32.2, we observed colocalization between snoring (index variant rs2664299) and volume of the left thalamus proper (index variant rs2664299). The PPH4 of Bayesian colocalization analysis is 0.997. We also observed the shared associations (LD *r*^2^ ≥ 0.6) with self-reported math ability, educational attainment, cognitive performance, and smoking initiation. **(C)** In 2q14.1, we observed colocalization between sleep duration (index variant rs62158206) and resting functional connectivity between the somatomotor and auditory networks (index variant rs62158170). The PPH4 of Bayesian colocalization analysis is 0.991. We also observed the shared associations with insomnia and diastolic blood pressure. **(D)** In 19q13.32, we observed colocalization between insomnia (index variant rs429358) and task functional connectivity in the secondary visual network (index variant rs429358). The PPH4 of Bayesian colocalization analysis is 0.98. We also observed the shared associations with Alzheimer’s disease, cognitive impairment, type 2 diabetes, myocardial infarction, and total cholesterol levels.

Five loci contributed to both sleep and resting/task fMRI traits. We found shared genetic influences between resting functional connectivity and sleep duration in the locus 2q14.1 (**Fig. 4C**). In addition, the locus 19q13.32 was associated with both insomnia and resting/task fMRI (**Figs. 4D** and **S19**). This locus harbors the *APOE* gene, a well-established genetic factor for Alzheimer’s disease. We also found that this locus had shared genetic influences for insomnia and the average proton density fat fraction in the liver (**Fig. S20**). Previous research has indicated an elevated risk of Alzheimer’s disease among individuals with NAFLD^114,115^ and sleep traits have also been consistently linked to Alzheimer’s disease and other dementias^2^. In addition, 11q13.2 contributed to both chronotype and fat fraction in the viable muscle tissue in the posterior thigh (**Fig. 5A**). Furthermore, we found five genetic loci that were associated with sleep and cardiac MRI traits, most of which were with chronotype. These loci encompass crucial genes associated with circadian rhythm regulation. For example, in 11p11.2, shared genetic influence was observed between the myocardial-wall thickness at end-diastole and both insomnia and chronotype (**Figs. 5B** and **S21**). This locus contains *CRY2,* a cryptochrome gene that is a critical component of the molecular clock and serves as a circadian photoreceptor^116^. Another locus, 22q13.1, contains the crucial circadian regulator *CSNK1E*^117^ and was associated with chronotype and the maximum area of the ascending aorta (**Fig. S22**).

**Fig. 5.**
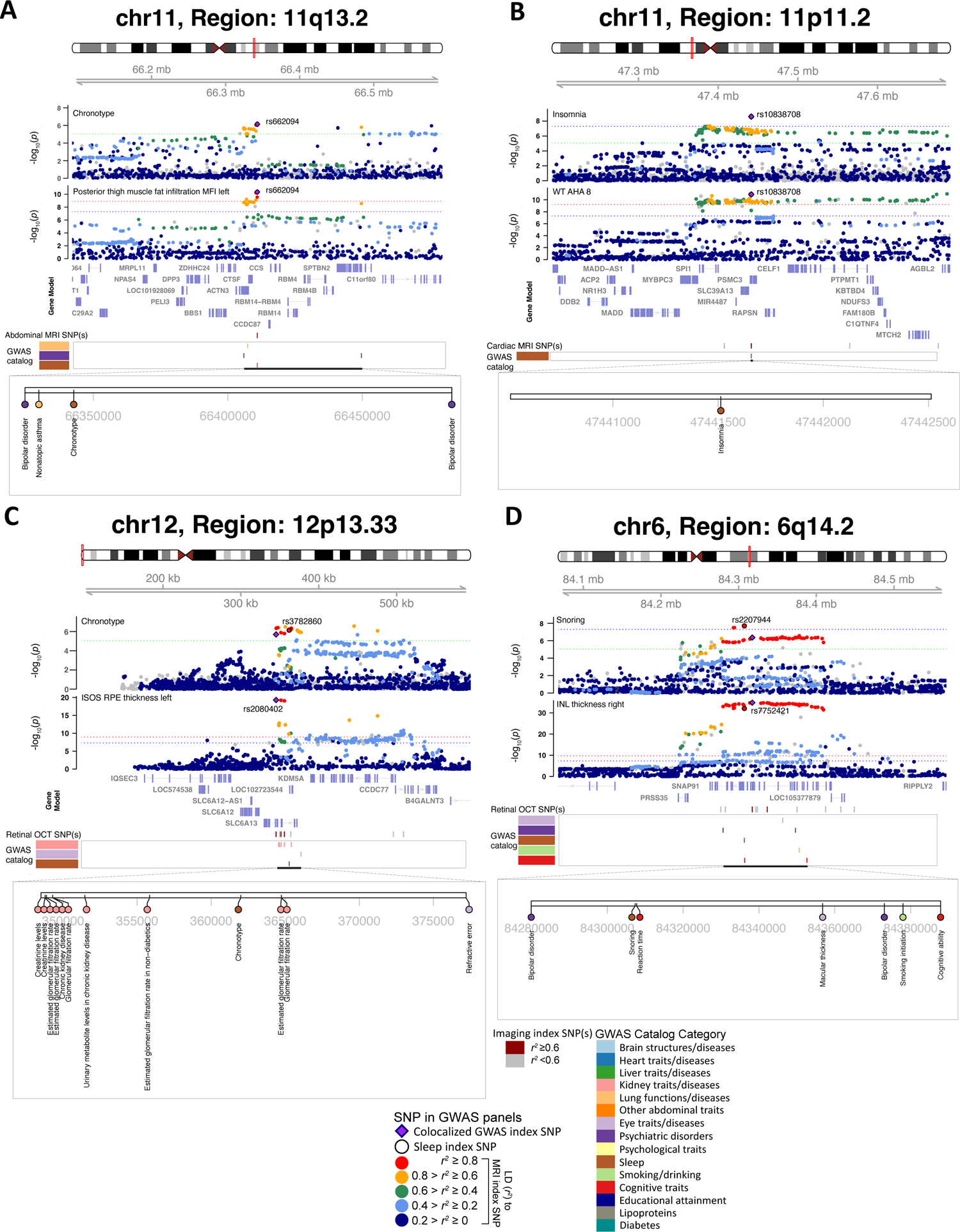
Selected genetic loci that were associated with both sleep and non-brain imaging traits. **(A)** In 11q13.2, we observed colocalization between chronotype (index variant rs662094) and the muscle fat infiltration in the left posterior thigh (index variant rs662094). The posterior probability of Bayesian colocalization analysis for the shared causal variant hypothesis (PPH4) is 0.986. We also observed the shared associations (LD *r*^2^ ≥ 0.6) with bipolar disorder and nonatopic asthma. **(B)** In 11p11.2, we observed colocalization between insomnia (index variant rs10838708) and regional myocardial-wall thickness at end-diastole (index variant rs10838708). The PPH4 of Bayesian colocalization analysis is 0.97. **(C)** In 12p13.33, we observed colocalization between chronotype (index variant rs3782860) and the average thickness measured between the inner and outer photoreceptor segments (ISOS) to the retinal pigment epithelium (RPE) across all subfields in the left eye (index variant rs2080402). The PPH4 of Bayesian colocalization analysis is 0.978. We also observed the shared associations with chronic kidney disease and estimated glomerular filtration rate. **(D)** In 6q14.2, we observed colocalization between snoring (index variant rs2207944) and the average inner nuclear layer (INL) thickness across all subfields in the right eye (index variant rs7752421). The PPH4 of Bayesian colocalization analysis is 0.883. We also observed the shared associations with bipolar disorder, smoking initiation, and cognitive ability.

We identified 17 loci that contributed to both sleep and retinal OCT traits, most of which were related to chronotype. The locus 14q24.3 was a key contributor to chronotype and a substantial portion (27 out of 46) of the OCT traits (**Fig. S23**). This locus contains the *VSX2* gene, which is critical to retina development^118^. In addition, in 12p13.33, we found shared genetic influences between chronotype and the average thickness between ISOS and RPE in the left eye (**Fig. 5C**). *CACNA2D4* gene in this region is related to retinal cone dystrophy^119^. In addition, the genetic locus 3p24.2, which includes a circadian clock gene *NR1D2*^120^, exhibited shared genetic influences with insomnia and disc diameter (**Fig. S24**). Moreover, in 1p13.2, rs6537747 influences both chronotype and average thickness between ISOS to the RPE (**Fig. S25**). This genetic variant was a brain eQTL of *CAPZA1*, which was associated with various eye-related phenotypes. It has been reported that knockout mice lacking *CAPZA1* display multiple eye diseases, such as cataract and abnormal retinal blood vessel morphology^121^. Genetic overlaps with snoring were also observed, such as with the average INL thickness in 6q14.2 (**Fig. 5D**).

### Sleep-disease genetic correlation patterns across multiple organs

Given the substantial sleep-imaging genetic links between sleep and organ structures and functions, we aimed to elucidate the genetic connections between sleep and relevant diseases of these organs from a multi-organ perspective. We systematically collected 113 GWAS datasets of 50 distinct diseases/traits spanning these organs, including 7 psychiatric/neurological disorders, 13 cardiovascular diseases/traits, 2 diabetes, 2 eye diseases, 10 kidney diseases/traits, 4 liver diseases/traits, and 12 lung functions/diseases (**Table S7**). At a 5% FDR level (*P* range = (8.85×10^-117^, 1.55×10^-2^)), an atlas of sleep-disease genetic correlations was then established by performing LD score regression^93^ with the 34 sleep GWAS summary statistics (**Table S8**).

Across all these organs, brain-related diseases had the strongest genetic correlations with multiple sleep traits (**Figs. 6A-6B** and **S26**). Specifically, neuroticism and depression had the highest genetic correlations with insomnia (β range = (0.38, 0.47), *P* < 1.13×10^-27^ for neuroticism and β range = (0.33, 0.60), *P* < 1.16×10^-6^ for depression, **Fig. S27**). Insomnia is frequently observed in individuals with depression^122^, and their sleep patterns exhibit a decrease in slow-wave sleep and an increase in both rapid eye movement (REM) density and total REM sleep time^123^. The close association between neuroticism and insomnia was also consistent with previous studies, with neuroticism being identified as one of the key factors contributing to inadequate sleep^124,125^. In addition to insomnia, we found that other sleep traits also had broad genetic correlations with mental disorders. For example, schizophrenia, bipolar disorder, depression, and neuroticism were genetically correlated with difficulty in getting up in the morning and narcolepsy (β range = (−0.35, 0.60), *P* < 1.07×10^-5^). Narcolepsy has been shown to exhibit a high comorbidity with psychiatric disorders^126^. In addition, being an evening person exhibited genetic correlation with an elevated risk of schizophrenia (β range = (−0.15, −0.09), *P* < 7.27×10^-6^), which was consistent with previous findings that evening chronotype is a risk factor for multiple mental health problems, including schizophrenia^127^. Moreover, schizophrenia consistently exhibited genetic correlations with most sleep traits we considered, except insomnia (β range = (−0.20, 0.16), *P* < 7.27×10^-6^). Significant genetic correlations were also observed with Alzheimer’s disease and Parkinson’s disease, both of which exhibited genetic correlations with shorter sleep durations (β range = (−0.20, 0.13), *P* < 8.20×10^-3^), matching findings from previous studies^2^ (**Figs. 6A** and **S27**).

**Fig. 6.**
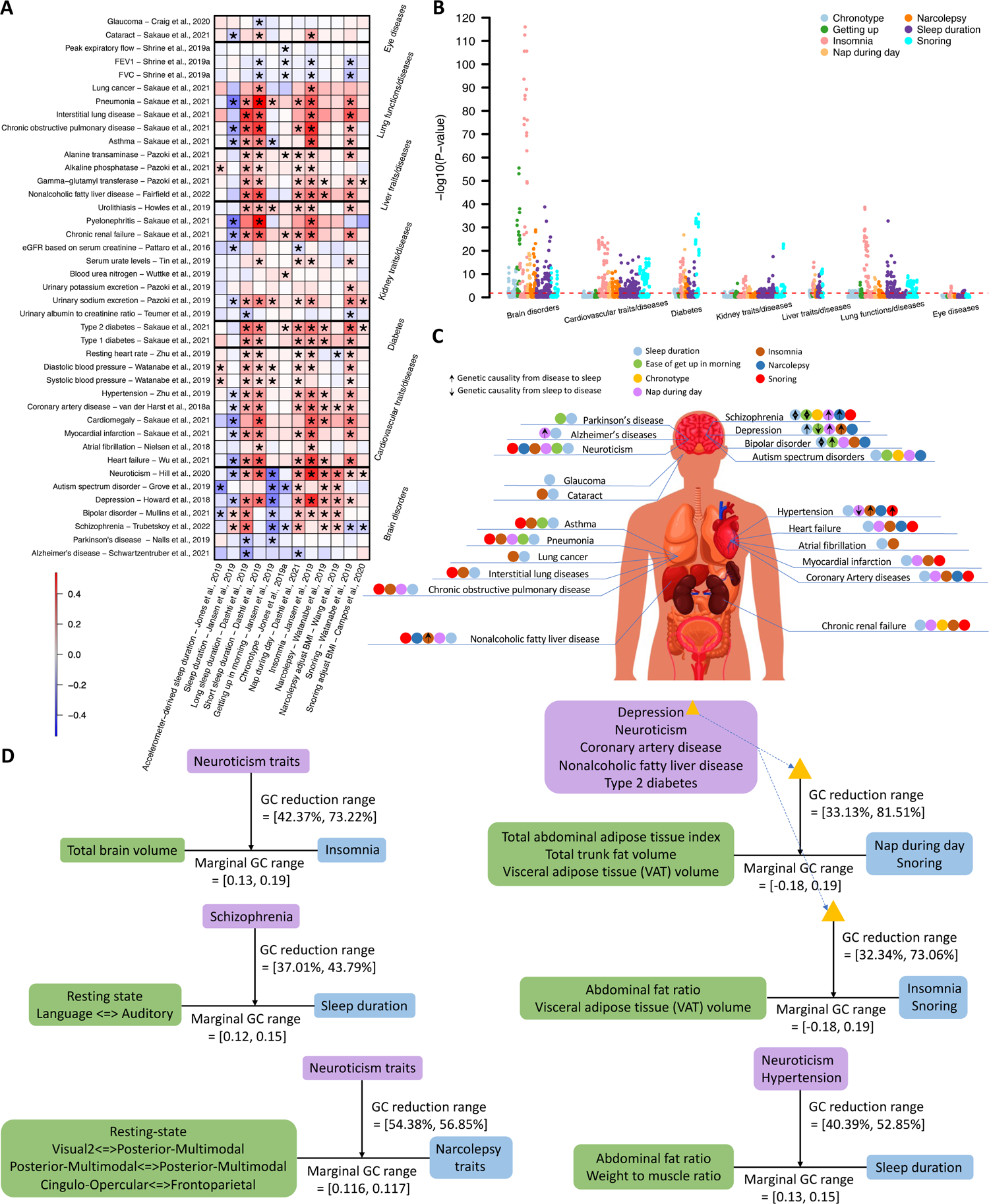
Genetic connections between sleep and multi-organ diseases/traits as well as the mediation effects of disease. **(A)** Selected genetic correlations between sleep traits (*x*-axis) and diseases (*y*-axis) across different organs. The color represents correlation estimates. Associations that passed a 5% FDR rate (*P* < 1.55×10^-2^) were marked with an asterisk. See **Tables S3** and **S7** for more information of sleep traits and diseases/traits, respectively. **(B)** −log10(P-value) between sleep and diseases/traits. Sleep traits were highlighted with different colors. A red dashed line represents the *P* value threshold from Bonferroni correction (*P* < 1.30×10^-5^), and a black dashed line represents *P* value threshold from a 5% FDR rate (*P* < 1.55×10^-2^). **(C)** Summary of genetic correlations and MR results between sleep and diseases/traits. A circle next to a disease/trait represents a genetic correlation between a sleep trait and the disease/trait. The colors within the circles represent different sleep traits. The arrows represent in MR results. Specifically, an upward arrow represents genetic causality from diseases/traits to sleep, whereas a downward arrow represents genetic causality from sleep to diseases/traits. Both upward and downward arrows represent bi-directional causality. **(D)** Summary of mediation effects of diseases on selected associations between imaging and sleep traits. The marginal GC represents the marginal genetic correlation between imaging and sleep traits. The GC reduction is calculated as the difference in genetic correlations between imaging and sleep traits with and without mediation by disease, divided by the marginal genetic correlation estimates.

Sleep and cardiovascular diseases have close connections^128,129^. We found that the majority of genetic correlations with cardiovascular diseases and traits were predominantly related to snoring, narcolepsy, insomnia, and sleep duration. Specifically, multiple heart diseases, such as heart failure, myocardial infarction, coronary artery disease, and hypertension, showed widespread genetic correlations with snoring, insomnia, and shorter or longer sleep duration (β range = (0.13, 0.40), *P* < 1.01×10^-5^).

These sleep traits also had genetic correlations with systolic/diastolic blood pressure and resting heart rate (β range = (0.05, 0.17), *P* < 1.41×10^-2^). Other sleep traits also had genetic correlations with a few specific heart conditions. For example, narcolepsy and a more frequent daytime nap were genetically correlated with heart failure, coronary artery disease, and hypertension (β range = (0.11, 0.18), *P* < 7.46×10^-6^).

Diseases related to the kidney, liver, and lung tended to have similar patterns in their genetic correlations with sleep (**Fig. S27**). For example, snoring, insomnia, and shorter or longer sleep duration were genetically correlated with increased risk of chronic renal failure, NAFLD, asthma, COPD, and pneumonia (β range = (0.11, 0.53), *P* < 1.12×10^-5^), as well as type 1 and type 2 diabetes (β range = (0.16, 0.34), *P* < 8.64×10^-6^). Consistent genetic correlations between sleep and the relevant biomarkers of these diseases were observed. For example, urinary biomarkers such as urinary potassium to creatinine ratio, urinary sodium to potassium ratio, and urinary sodium excretion, along with liver biomarkers such as liver enzyme levels, displayed genetic correlations with snoring, insomnia, and shorter or longer sleep duration as well (β range = (−0.38, 0.27), *P* < 1.19×10^-5^). Furthermore, higher forced expiratory volume (FVC) was genetically correlated with a reduced risk of snoring (β range = (−0.13, −0.10), *P* < 2.74×10^-6^). Significant genetic correlations were also observed with eye diseases. Cataract was genetically correlated with shorter sleep duration (β range = (0.12, 0.16), *P* < 1×10^-2^) and increased risk of insomnia (β range = (0.10, 0.20), *P* < 1.43×10^-2^). Cataracts limit the transmission of light to the area of the eye responsible for regulating the biological clock, which might cause insomnia. Cataract patients with sleep problems showed improved sleep quality following cataract surgery^130,131^.

Beyond the above organ-specific patterns, we identified distinct genetic correlation patterns across various organs for different sleep traits (**Fig. S27**). For example, the majority of genetic correlations with chronotype and ease of waking up were associated with brain disorders, rather than diseases of other organs. This was different from other sleep traits, which typically had a more balanced pattern of associations across various organs. In addition, after adjusting for BMI, the correlations between snoring and narcolepsy with many heart, kidney, liver, and lung diseases diminished, but their genetic links with mental disorders, such as schizophrenia, remained. This suggests that BMI may play a distinct role in the genetic correlations between snoring/narcolepsy and these non-brain organs compared to its role in sleep’s correlations with psychiatric disorders. Furthermore, evidence of nonlinear genetic relationships between sleep duration and various diseases was observed across all organs. This was deduced from results showing both excessively short and long sleep durations, as well as insomnia, having consistently similar positive genetic correlations with various diseases, while regular sleep duration traits typically showed negative correlations. These observations suggest that both short and long sleep durations may adversely affect overall brain and body health.

Using 23 sets of publicly available disease GWAS summary statistics from the East Asian population (**Table S9**), we further explored their cross-ancestry sleep-disease genetic correlations with sleep using Popcorn^132^ (**Methods**). Compared to genetic correlations within the European ancestry, the direction of cross-ancestry genetic correlations remained largely consistent for several diseases, though the magnitude of estimates was lower (**Figs. S28-S32** and **Table S10**). For example, of the 25 pairs demonstrating significant genetic correlations between sleep and type 2 diabetes within the European population, 24 maintained consistent directions in the cross-population analysis between East Asians and Europeans (**Fig. S28**). Furthermore, the gap in the two sets of correlation estimates might be partly attributed to the different genetic architectures of the same disease across the two populations. For example, the average reduction in genetic correlations between European sleep traits and East Asian type 1 diabetes, compared to the within-European correlations, was 0.38 (**Fig. S29**). Meanwhile, the genetic correlation between European and East Asian type 1 diabetes was 0.56, indicating that a large portion of the genetic correlation reduction may come from the genetic differences in type 1 diabetes between the two populations. These observations may imply that the sleep-disease genetic correlation patterns also exist in other populations, with similar directions and comparable magnitudes of correlation.

Furthermore, genetic causal links between sleep and diseases/traits were explored using two-sample Mendelian randomization (MR) with 44 sets of disease GWAS summary statistics, ensuring no overlap with subjects from the sleep GWAS (**Table S11** and **Methods**). At a 5% FDR level (*P* < 2.93×10^-3^), we found evidence of causal links between sleep and diseases in both directions, predominantly involving psychiatric disorders (**Fig. 6C** and **Table S12**). Among all studied diseases, the strongest causal effects were observed from multiple psychiatric disorders on sleep traits. Specifically, we found consistent evidence that depression caused an increased risk of insomnia (*P* < 7.12×10^-6^, **Fig. S33**). We also found causal links between schizophrenia and sleep, where schizophrenia was causally associated with longer sleep duration (*P* < 9.66×10^-8^) and difficulty in getting up in the morning (*P* < 1.84×10^-6^, **Fig. S34**). Similar to schizophrenia, bipolar also caused longer sleep duration (*P* < 1.11×10^-3^, **Fig. S35**) and difficulty in getting up in the morning (*P* < 8.94×10^-4^, **Fig. S36**). These findings were generally consistent among multiple sleep GWAS summary statistics data resources. Among brain disorders, we also found that Alzheimer’s disease decreased the genetic liability of daytime napping (*P* < 1.32×10^-3^, **Fig. S37**). Moreover, two non-brain disorders, NAFLD and hypertension, had causal effects on sleep. For example, we found that NAFLD increased the risk of insomnia (*P* < 5.83×10^-11^, **Fig. S38**) and there was a causal effect from hypertension to snoring (*P* < 1.76×10^-3^, **Fig. S39**). These results provide further insights into the genetic connections between sleep and both brain and body health, highlighting the notably intricate causal links between sleep and psychiatric disorders, as evidenced by the currently available data.

### Diseases genetically involved in sleep-imaging connections

After mapping both the multi-organ sleep-imaging and sleep-disease genetic correlations, we would like to explore whether diseases mediate the sleep and imaging connections, potentially offering new insights into the complex interrelations among sleep, imaging, and health conditions. We first identified 623 combinations of sleep, imaging, and disease triples that had pairwise marginal genetic correlations, and then we analyzed the mediation effects using genomic structural equation modeling^133^ (**Methods**). We found 145 sleep-imaging pairs whose associations were mediated by the presence of disease at a 5% FDR level, most (133/145) of which exhibited consistent directions in marginal and conditional genetic correlations (**Figs. 6D** and **S40, Table 13**). Below we highlight several patterns we found, particularly for brain and abdominal imaging traits.

There were 12 pairs of genetic correlations between brain MRI traits and sleep traits mediated by psychiatric disorders, specifically schizophrenia and neuroticism (**Fig. S41**). Schizophrenia was frequently involved in the genetic correlations between sleep and resting fMRI traits. For example, schizophrenia mediated the association between functional connectivity between language and auditory networks and sleep duration (mediation proportion range = [37.01%, 43.79%]), as well as the association between functional connectivity between cingulo-opercular and default mode networks and narcolepsy (mediation proportion range = [44.00%, 50.75%]). In addition, neuroticism played a role in mediating the association between functional connectivity in the posterior-multimodal network and narcolepsy (mediation proportion = 54.38%), as well as the association between total brain volume and insomnia (mediation proportion range = [42.37%, 73.22%]).

The genetic correlations between abdominal MRI traits and sleep were mediated by various diseases, which constituted a large majority of the observed pairs (94 out of 133, **Fig. S42**). Particularly, links between abdominal MRI and insomnia, daytime napping, and snoring were consistently mediated by a common set of diseases spanning multiple organs, including depression, neuroticism, coronary artery disease, NAFLD, and type 2 diabetes. These diseases were widely involved in genetic correlations between daytime napping and liver fat percentage, total abdominal adipose tissue index, total trunk fat volume, and VAT (mediation proportion range = [34.92%, 76.84%]). The same set of diseases also mediated the associations between insomnia and weight-to-muscle, abdominal fat ratio, total and posterior thigh fat free muscle volumes, and VAT (mediation proportion range = [32.34%, 97.86%]). Notably, the association between weight-to-muscle ratio and insomnia was substantially mediated by depression, resulting in the highest reduction in genetic correlation observed with and without considering the mediation effect (mediation proportion = 97.86%). Additionally, these diseases mediated the associations between snoring and total abdominal adipose tissue index, total trunk fat volume, abdominal fat ratio, pancreas fat ratio, subcutaneous fat volume, and VAT (mediation proportion reduction range = [33.13%, 89.33%]). These results provide additional insights into the genetic connections between sleep and imaging traits, which potentially reflect the involved genetic risk factors for related diseases.

## DISCUSSION

Sleep is vital to both physical health and mental well-being. Understanding how sleep interacts with human health is of great interest. This study aimed to map phenotypic and genetic connections between sleep and whole-body health by using multi-organ imaging data as endophenotypes. Our large-scale datasets and analyses uncovered substantial links between sleep and the structure and function of various organs, revealing organ-specific patterns for different sleep traits. We examined the genetic basis of sleep-imaging connections from different perspectives, investigating both the shared genetic loci and the overall genetic similarity across the genome. Some genetic loci also contributed to both sleep and diseases related to these studied organs. Significant genetic correlations were identified between sleep and a wide spectrum of diseases, especially neurological and psychiatric disorders, cardiovascular diseases, and chronic kidney and liver diseases. Consistent with previous findings^45,134^, we found causal links with multiple psychiatric disorders, such as depression, schizophrenia, and bipolar disorders. Future epidemiological and clinical studies of sleep and comorbid conditions may benefit from our results and insights.

In this study, we identified many novel genetic loci that were associated with both sleep and imaging traits. Several genetic loci exhibited pleiotropy between sleep and imaging traits across various organs, including 17q21.31, 4q24, and 5q14.3. Additionally, numerous other loci appeared to be specific to particular organs (**Fig. 3B**). We found that chronotype and insomnia played a predominant role in these genetic effects shared between sleep and organ structures and functions, which may be related to the extensive involvement of the loci in the regulation of circadian rhythms^135^. Genes associated with circadian rhythms were intricately linked to the underlying mechanisms that govern fundamental biological processes for human health, spanning multiple organs^50^. The shared genetic influences with sleep extended across a wide variety of imaging modalities, with the highest number of loci being associated with brain white matter, brain grey matter, and eye-related traits. Notably, the genetic loci associated with these imaging modalities often vary, underscoring the value of a multi-organ, multi-modality approach to mapping sleep’s connections to human health. Interestingly, we did not find shared genetic loci between narcolepsy and any of the imaging data categories. Narcolepsy, being an autoimmune disease, is strongly influenced by both environmental and genetic factors^136^. The limited presence of narcolepsy in the genetic pleiotropy with imaging traits could be attributed, in part, to the relatively low statistical power of current narcolepsy GWAS. This is partially supported by the robust genome-wide genetic correlation findings between narcolepsy and resting MRI traits, which did not require the identification of specific GWAS significant loci (**Fig. 2B**).

Psychiatric disorders exhibited robust genetic connections with sleep in different approaches. We found that several genetic loci affecting both sleep and imaging traits were also linked to psychiatric disorders, such as 9q22.31 and 18q21.2 that were associated with depression (**Figs. S12** and **S18**). Genetic correlations were also found between sleep and various psychiatric disorders. Different from many diseases related to non-brain organs, most genetic correlations between psychiatric disorders and sleep still existed after adjusting for BMI. Additionally, MR revealed causal genetic effects from psychiatric disorders, like depression and schizophrenia, on sleep patterns. Some of the causal effect estimates survived several sensitivity analyses and yielded similar effect sizes, supporting the robustness of the causal links between mental disorders and sleep. These collective findings suggest that psychiatric disorders distinctly stand out and have a notably close relationship with sleep, when viewed from a whole-body perspective across various diseases.

We identified many pairs of imaging and sleep traits whose genetic correlations were mediated by diseases (**Fig. 6D**). For example, psychiatric disorders such as schizophrenia and neuroticism mediated the genetic link between resting fMRI and narcolepsy, as well as between total brain volume and insomnia. Additionally, diseases affecting multiple body systems played a role in the genetic connections between abdominal MRI and sleep traits, including depression, neuroticism, coronary artery disease, NAFLD, and type 2 diabetes. It is worth mentioning that our genetic mediation analysis alone did not imply causal directions but rather suggested the diseases involved in the identified sleep-imaging genetic correlations. It has been reported that brain structures tend to affect narcolepsy, as the loss of neurons producing hypocretin, controlled by specific genes, is the main cause of narcolepsy^137^. Conversely, insomnia could be either a cause or a consequence of brain structural changes^138^. The relationship between body fat/muscle composition and visceral fat with sleep may also be bidirectional. Circadian misalignment and daytime napping elevate cortisol levels, potentially leading to fat accumulation in the abdominal area^139,140^. Abdominal fat affects airway size and function, which may cause OSA, and OSA contributes to obesity due to lack of sleep, daytime drowsiness, and metabolic disturbances^141^. Regardless of causal directions, the identified mediated effects aid in understanding the genetic risks of diseases underlying sleep-imaging connections.

The present study has several limitations. First, our sleep-imaging and sleep-disease analyses were mainly based on European subjects. We conducted a cross-ancestry genetic correlation analysis using East Asian disease GWAS summary statistics, which indicated a level of similarity in sleep-disease connections between these two populations. In future studies, data from a broader range of sources could be incorporated into the imaging genetics study of sleep, and findings identified by European cohorts may be generalized to global populations. The current study focused on the brain, heart, eye, and abdominal organs. Among them, detailed imaging measurements were extensively conducted for the brain and heart. However, the imaging analysis of the liver, kidney, lung, spleen, and pancreas was less comprehensive. Improving abdominal MRI pipelines and incorporating data from additional organs could provide further insights into sleep research. In addition, due to the nature of the data resources, the sleep traits and imaging data used in this study were not collected at the same time point. Finally, we focused primarily on linear relationships in our models. Some of our results and several recent studies have demonstrated that certain sleep traits may also have nonlinear phenotypic and genetic links (such as sleep duration^31,37^). In the future, advanced statistical and machine learning methods in imaging genetics may be used to identify more comprehensive relationships.

## METHODS

Methods are available in the ***Methods*** section. *Note: One supplementary information pdf file and one supplementary table zip file are available*.

## Data Availability

The GWAS summary statistics for brain MRI, cardiac MRI, as well as for derived OCT traits, can be accessed from https://bigkp.org/, https://heartkp.org/, and https://eyekp.org/, respectively. The GWAS summary statistics for the abdominal MRI traits generated in our study will be publicly released upon publication. GWAS summary statistics of sleep and diseases used in this study are publicly available, and the details can be found in the Supplementary Tables and the Supplementary Note of this study. The individual-level UK Biobank data used in this study can be obtained from https://www.ukbiobank.ac.uk/.

## ACKNOWLEDGEMENTS

We thank the individuals represented in the UK Biobank study for their participation and the research teams for their work in collecting, processing, and disseminating these datasets for analysis. We would like to thank the research computing groups at the University of North Carolina at Chapel Hill, Purdue University, and the Wharton School of the University of Pennsylvania for providing computational resources and support that have contributed to these research results. We gratefully acknowledge all the studies and databases that made GWAS and eQTL summary-level data publicly available. This research has been conducted using the UK Biobank resource (application number 76139), subject to a data transfer agreement. Research reported in this publication was supported by the National Institute On Aging of the National Institutes of Health under Award Number RF1AG082938. The content is solely the responsibility of the authors and does not necessarily represent the official views of the National Institutes of Health. The study has also been partially supported by funding from the Purdue University Statistics Department and the Department of Statistics and Data Science at the University of Pennsylvania. Assistance for this project was also provided by the UNC Intellectual and Developmental Disabilities Research Center (NICHD; P50 HD103573).

## AUTHOR CONTRIBUTIONS

B.Z. designed the study. Z.F., Y.Y., Y.G., Y.L., J.S., X.Y., B.L., J.L., and T.L. analyzed the data. Q.W., C.G., P.P., T.L., P.G., and H.Z. provide comments and interpret the results. Z.F. and B.Z. wrote the manuscript with feedback from all authors.

## COMPETING FINANCIAL INTERESTS

The authors declare no competing financial interests.

## METHODS

### Phenotypic sleep-imaging analyses

The UKB study (https://www.ukbiobank.ac.uk/) recruited approximately half a million participants aged between 40 and 69 years between 2006 and 2010^71^. The ethics approval of the UKB study was from the North West Multicentre Research Ethics Committee (approval number: 11/NW/0382). Here we examined the phenotypic associations between sleep and organs using multi-modal MRI measurements of the brain, heart, and abdomen, as well as derived OCT traits of the retina. Detailed procedures to generate brain and cardiac MRI traits used in our analyses can be found in previous papers^65,67,69,72,142^. All these imaging traits were generated from the raw images in Category 100003 (https://biobank.ndph.ox.ac.uk/showcase/label.cgi?id=100003). Within the brain, three major brain MRI modalities were examined: 1) brain anatomical and neuropathological structures from structural MRI, including regional brain volumes^65^ and cortical thickness traits^72^; 2) white matter microstructures from diffusion MRI, including multiple DTI parameters^67^; and 3) intrinsic and extrinsic functional organizations of the cerebral cortex from resting and task fMRI^69^. Regarding the heart, we used 82 traits extracted from short-axis, long-axis, and aortic cine cardiac MRI^72^, including global and regional measures of 4 cardiac chambers (the left ventricle, right ventricle, left atrium, and right atrium), as well as 2 aortic sections (the ascending aorta and descending aorta). Abdominal imaging traits comprised MRI data of the liver, kidneys, lungs, pancreas, spleen, and body muscle/fat composition^22–24,73–79^. For the eyes, derived OCT measures were used, providing measurements into the thickness of different retinal layers and their subfields, as well as the vertical cup-to-disc ratio and disc diameter^70^. Detailed information regarding the collection and processing of imaging data, muti-organ diseases/traits, and sleep data can be found in the **Supplementary Note**.

In the phenotypic analysis, we used the unrelated white British imaging individuals in UKB phases 1 to 3 data release as the discovery sample (mean age range = (45,82), mean = 64.15, standard error = 7.67, and proportion of females was 51.6%). We fitted linear models for each pair of sleep and imaging traits (average *n* = 35,893 for self-reported sleep traits, average *n* = 15,337 for accelerometer-derived sleep traits, **Table S2**). We adjusted for the basic set of covariates, including age (at assessment and imaging), age squared, sex, age-sex interaction, age-squared-sex interaction, assessment center code, imaging site code, top 10 genetic principal components (PCs)^143^, BMI, and Townsend deprivation index (TDI). For device-measured traits, we also adjusted for age at accelerometry measurement and season of wearing the device. Previous UKB studies on sleep or physical activity traits frequently accounted for these basic covariates, such as assessment center^46,51^, BMI^144–146^, TDI^144,146,147^, and season of wearing the device^148^. For all brain MRI traits, we adjusted for volumetric scaling, head motion, head motion-squared, brain position, and brain position-squared^64,66^. For regional brain volumes and regional cortical thickness measures, we additionally adjusted for the total brain volume and global mean thickness, respectively, to remove the global effects. For abdominal MRI traits, we adjusted for standing height. For cardiac MRI traits, we adjusted for standing height, standing height to the 1.7th power, standing height to the 2.1th power, standing height cubed, weight, body surface area to the 1.5th power, pulse rate, FVC, forced expiratory volume in the first second, preserved ejection fraction, peripheral and central pulse pressure during pulse wave analysis, average heart rate, body surface area, whole body fat-free mass, pulse wave arterial stiffness index, and waist-to-hip ratio^72^. The values greater than five times the median absolute deviation from the median were removed in each trait and continuous covariate variable. The *P* values from the two-sided *t* test were reported (R version 3.6.0) and we controlled a 5% false discovery rate (FDR) using the Benjamini-Hochberg procedure.

To evaluate the robustness of these results, we used two approaches. In both approaches, we examined whether the significant associations identified in the discovery analysis had the same effect direction and had a *P* value less than 0.05. The first one was to repeat the analysis using a small hold-out set of data from non-white British UKB subjects (average *n* = 3,955 for self-reported sleep traits and average *n* = 1,383 for accelerometer-derived sleep traits). The second was to adjust a more comprehensive set of covariates on the same discovery sample (that is, unrelated white British subjects). For brain MRI traits, we additionally incorporated covariates related to diet (e.g. vegetable and meat intake), socioeconomics (e.g., education, employment status), bone minerals, overall fitness (e.g. disability), cognitive functions, social support (e.g. leisure/social activity frequency), family disease history, diagnosed medical conditions from both questionnaire and ICD-10 (e.g. cancer, diabetes). For non-brain imaging traits, covariates related to cognitive functions and social support were excluded as they contained too many missing values and exhibited limited relevance to these imaging traits. This broader range of covariates was established by referencing many previous human health studies on sleep and other physical activity traits. For example, studies examining the association between brain structures, dementia, and sleep highlighted the relevance of socioeconomic factors, such as education level and employment status, in analyzing connections with brain-related traits^144–147,149^. Similarly, other research has demonstrated significant associations with bone minerals^20^, body fat percentage^150^, and cancer risk^151,152^, which motivated us to also include these body fitness and disease variables as covariates.

### Genetic sleep-imaging analyses

Our genetic analyses were mainly based on GWAS summary statistics of sleep and imaging traits. We systematically looked for the genomic loci that were reported to be significant for both sleep and imaging traits. First, we obtained the reported imaging-associated variants from previous studies, including 101 regional brain volumes^65^, 63 cortical thickness^72^, 110 DTI parameters^67^, 1,999 network-level parcellation-based resting and task fMRI traits^69^ (1,080 for resting and 919 for task), 82 cardiac MRI traits^72^, and 46 derived OCT traits^70^. Bonferroni correction has been used in each of these studies to control for the number of imaging traits analyzed, which resulted in imaging modality-specific *P* value thresholds used in these studies, including *P* < 4.55×10^-10^ for regional brain volumes, *P* < 7.94×10^-10^ for cortical thickness, *P* < 2.32×10^-10^ for DTI parameters, *P* < 4.63×10^-11^ for resting fMRI, *P* < 5.44×10^-11^ for task fMRI, *P* < 6.10×10^-10^ for cardiac MRI, and *P* < 1.09×10^-9^ for derived OCT measures.

Second, we performed GWAS for the 41 abdominal MRI traits using linear mixed effect models via fastGWA^153^. We used the imputed genotyping data from the UKB study, adhering to the standard quality controls used in previous studies^72^. These included 1) excluding individuals with a missing genotype rate greater than 0.1; 2) removing variants with a missing genotype rate over 0.1; 3) discarding variants with a minor allele frequency (MAF) less than 0.01; 4) eliminating variants that failed the Hardy-Weinberg equilibrium test at the 1×10^-7^ level; and 5) omitting variants with an imputation INFO score below 0.8. We adjusted for the effects of standing height, BMI, age, sex, assessment center, age-squared, age-sex interaction, age-squared-sex interaction, and top 40 genetic PCs^143^. We used the *P* < 1.22×10^-9^ (that is, 5×10^-8^/41) as the GWAS significance threshold for abdominal MRI and defined the independent (LD *r*^2^ < 0.1) significant genetic loci using FUMA^154^ (version v1.3.8).

Next, among these imaging-associated variants (and variants in LD with them, *r*^2^ ≥ 0.6), we searched for those that were also reported in the NHGRI-EBI GWAS Catalog^103^ (https://www.ebi.ac.uk/gwas/, version 2022-07-09) to be significant in previous GWAS of various sleep traits. For obtained sleep-significant genetic variants, we further searched to see whether they were reported brain eQTLs on MetaBrain (https://www.metabrain.nl/)^189^. Finally, we tested for whether the sleep and imaging traits had shared causal genetic variants using the Bayesian colocalization test^130^ with default settings.

We examined genetic correlations between sleep and imaging traits via LD score regression^93^ (https://github.com/bulik/ldsc/, version 1.0.1). We collected and used 34 sets of publicly available GWAS summary statistics of sleep traits (**Table S3**). For imaging traits, we screened the 101 regional brain volumes^65^, 63 cortical thickness^72^, 110 DTI parameters^67^, 90 network-level parcellation-based resting and task fMRI traits, respectively^69^, 82 cardiac MRI traits^72^, 41 abdominal MRI traits^22–24,73–79^, and 46 derived OCT measures^70^. To evaluate the genetic correlations between sleep and diseases associated with these organs, we systematically collected 113 GWAS summary statistics of 50 diseases/traits span a wide range of organs, covering 7 neurological/psychiatric disorders, 13 cardiovascular diseases/traits, 2 diabetes, 2 eye diseases, 10 kidney diseases/traits, 4 liver diseases/traits, and 12 lung diseases/traits (See **Table S7** for a full list of the 113 GWAS summary statistics and **Supplementary Note** for more details).We screened them with the 34 sets of sleep GWAS summary statistics. We used the default setups of LDSC, in which the HapMap3^155^ variants and the variants in the major histocompatibility complex region were removed, and the LD scores were calculated from the 1000 Genomes data.

Cross-population genetic correlation was analyzed using Popcorn^132^. With the 34 publicly available sleep GWAS summary statistics from the European ancestry (Table S3), we analyzed cross-population genetic correlations with 23 diseases/traits from the East Asian ancestry (Table S9). We used the recommended regression-based estimator and the pre-computed scores provided by the authors. These scores were based on 1000 Genome EUR and EAS as reference panels for the European and East Asian populations, respectively.

MR analyses were performed with the R package TwoSampleMR^156^ (https://mrcieu.github.io/TwoSampleMR/) with various methods. Inverse variance weighted was used as the main analysis, and MR Egger, simple mode, weighted mode, and weighted median^157^ were used as sensitivity analyses. Our bidirectional MR analyses were performed using published GWAS summary statistics between pairs of sleep and diseases/traits that showed evidence of genetic correlations in the LD score regression analysis at a 5% FDR level. We used 34 publicly available sleep GWAS summary statistics with samples from the UK Biobank, which was the same as in the genetic correlation analysis. GWAS of diseases/traits that had no overlapping sample with UKB were used (see **Table S11** for the full list of diseases/traits GWAS used in the MR analyses). To select independent strong genetic instrumental variables, exposure GWAS summary statistics were clumped with the *P* value significance threshold being 5×10^-8^. The physical distance threshold for clumping was kb = 10,000, the LD threshold for clumping was *r*^2^ = 0.01, and we used the 1000 Genomes European reference panel. Harmonization was performed to ensure the effects of each genetic variant on exposure and outcome correspond to each other.

We conducted mediation analyses using the R package GenomicSEM^133^ (https://github.com/GenomicSEM/GenomicSEM). We selected 623 imaging-disease-sleep combinations that exhibited pairwise significant genetic correlations at a 5% FDR level and subsequently performed mediation analyses on these chosen combinations. For each combination, we fitted models of imaging and sleep traits both with and without mediation by disease. We reported the combinations where the genetic correlations between imaging and sleep traits were no longer significant after mediation by disease at a 5% FDR level. See **Table S13** for a full list of the 623 combinations of imaging, disease, and sleep traits that were selected for further mediation analyses, as well as the mediation analyses results.

## Code availability

We made use of publicly available software and tools. The codes are available upon reasonable request.

## Notes

### Competing Interest Statement

The authors have declared no competing interest.

### Author Declarations

We use the data from the UK Biobank (UKB). The data can be obtained from https://www.ukbiobank.ac.uk/. The ethics approval of the UKB study was from the North West Multicentre Research Ethics Committee (approval number: 11/NW/0382).

### Summary of Updates

Including more imaging and disease data resources to cover more organs and associated disorders.

