## Supplementary material for "The role of sleep in the human brain and body: insights from multi-organ imaging genetics": supp_information

**This PDF file includes:**

Supplementary Text
Supplementary Figures Figs. S1-S42
Legends for Tables S1 to S13

**Other Supplementary Materials for this manuscript include the following:**

Tables S1 to S13 (.xlsx) (available in a zip file)

### Supplementary Text

#### Imaging data types and processing.

Overall, we used 101 regional brain volumes and 63 cortical thickness measures from T1-weighted structural MRI[1-3], 110 DTI parameters from diffusion MRI[4], 90 parcellation-based network-level traits in resting and task fMRI[5], respectively, 46 derived OCT measures[6], 41 abdominal imaging traits, as well as 82 cardiac MRI traits from the short-axis, long-axis, and aortic cine images[3]. These traits captured the structural and functional characteristics of the human brain, abdomen, eye, heart, and aorta.

Briefly, the advanced normalization tools[7] (ANTs) was used to generate regional brain volumes for 98 cortical and subcortical areas, as well as 3 global brain volume measures, including the total gray matter volume, total white matter volume, and total brain volume. Similarly, 63 global and regional cortical thickness measures were also generated by ANTs. In addition, we applied the ENIGMA-DTI pipeline[8,9] to diffusion MRI and generated 110 tract-averaged parameters, including the fractional anisotropy (FA), mean diffusivity (MD), axial diffusivity (AD), radial diffusivity (RD), and mode of anisotropy (MO) for 21 predefined major white matter tracts, as well as across the whole brain (5 × 22). For resting and task fMRI, we used a parcellation-based approach with the Glasser360 atlas[10] and partitioned the cerebral cortex into 360 regions in 12 functional networks[11], including the primary visual, secondary visual, auditory, somatomotor, cingulo-opercular, default mode, dorsal attention, frontoparietal, language, posterior multimodal, ventral multimodal, and orbito-affective networks. We calculated the mean functional connectivity for each pair of networks (including within the same network) and mean functional connectivity of the whole cortex. Derived OCT measures contain measurements for thickness of different retinal layers and their subfields, as well as the vertical cup-to-disc ratio and disc diameter, which were derived from Category 100079 in the UK Biobank. Quality control was done as the suggestions outlined in Data-Fields 28552 and 2855343, retaining images with an image quality score > 45. Subsequently, only OCT measures with sample size greater than 30,000 were retained, resulting in 46 measures with an average sample size of 62,425.

Abdominal imaging traits were derived from Category 158[12,13], Category 126[14,15], Category 149[16-20], and Category 159[21] within the UK Biobank, encompass various measurements related to body fat/muscle composition, as well as characteristics of the liver, lung, kidney, pancreas, and spleen. To ensure data quality, images of low quality were removed, and identified through error indicators specified in Data-Fields 23363, 23358, 23359, 23361, 23364, 23360, 23362, and 23357. Outliers deviating more than ±5 median absolute deviations from the median were removed. Then, a rank-based inverse normal transformation was applied to achieve normality. These processes yielded 41 abdominal measures with an average sample size of 43,516. The 82 cardiac MRI traits were from 6 categories, including 64 traits of left ventricle, 4 of left atrium, 4 of right ventricle, 4 of right atrium, 3 of ascending aorta, and 3 of descending aorta. In total, there were 623 imaging traits across 8 imaging modalities (**Table S1**).

#### Sleep data in the phenotypic and genetic connection analyses.

For phenotypic association analyses, we studied 7 self-reported sleep traits and 3 accelerometer-derived sleep duration traits. The 7 self-reported sleep traits including sleep duration ("*About how many hours sleep do you get in every 24 hours? (please include naps)*", Data-Field 1160); getting up in morning *("On an average day, how easy do you find getting up in the morning? 1) not at all easy; 2) not very easy; 3) fairly easy; 4) very easy"*, Data-Field 1170); morning/evening person (chronotype, *"Do you consider yourself to be 1) definitely a 'morning' person; 2) more a 'morning' person than an 'evening' person; 3) more an 'evening' person than a 'morning' person; 4) definitely an 'evening' person?",* Data-Field 1180); nap during day ("*Do you have a nap during the day? 1) never/rarely; 2) sometimes; 3) usually.",* Data-Field 1190); sleeplessness/insomnia ("*Do you have trouble falling asleep at night or do you wake up in the middle of the night? 1) never/rarely; 2) sometimes; 3) usually.",* Data-Field 1200); snoring ("*Does your partner or a close relative or friend complain about your snoring?*", Data-Field 1210); and daytime dozing/sleeping (narcolepsy, "*How likely are you to doze off or fall asleep during the daytime when you don't mean to? (e.g. when working, reading, or driving) 1) never/rarely; 2) sometimes; 3) often; 4) all of the time.",* Data-Field 1220). We used the data coded by the UKB study and removed the subjects with responses "do not know" or "prefer not to answer". The 3 accelerometer-derived sleep duration measurements were collected from Return 1942, Return 2242, and Data-Field 40046 across multiple studies[22-24]. Subjects with inadequate wear time (Data-Field 90015), poor calibration (Data-Field 90016), or missing calibration (Data-Field 90017) were removed.

For genetic correlation analyses, we used 34 publicly available sleep genome-wide association studies (GWAS), covering 7 distinct sleep traits. These contained 5 GWAS on self-reported sleep duration[25-27] (including 1 short sleep duration and 1 long sleep duration), 5 accelerometer-derived sleep duration[22,28,29] (including 1 short sleep duration and 1 long sleep duration), 2 on ease of getting up in the morning[25,26], 6 on chronotype[25-27,30,31], 2 on daytime napping[26,32], 5 on insomnia[25,26,33-35], 4 on narcolepsy[25,26,36] (including 1 adjusted for BMI), and 5 on snoring[25,26,37,38] (including 1 adjusted for BMI). All sleep GWAS were on human reference GRCh37 (hg19). See **Table S3** for a full list of the sleep GWAS used.

**Clinical outcomes collection and quality control.**

To evaluate the genetic similarities between sleep and diseases associated with these organs, we systematically collected 113 GWAS summary statistics of 50 diseases/traits spanning a wide range of organs, covering 7 neurological/psychiatric disorders, 13 cardiovascular diseases/traits, 2 diabetes, 2 eye diseases, 10 kidney diseases/traits, 4 liver diseases/traits, and 12 lung diseases/traits. We aimed to comprise a diverse set of diseases/traits and collected datasets that had the largest sample sizes. These were obtained from various sources, including knowledge portals like the Human Genetics Amplifier and GWAS Catalog[39]. Disease-specific consortia, such as CKDGen[40] for kidney-related traits, were also used. GWAS from the East Asian ancestry for cross-population genetic correlation analyses were mainly collected from Sakaue et al[41]. Additionally, GWAS originally based on the human reference GRCh38 (hg18) were liftover to GRCh37 (hg19) to ensure alignment with other GWAS datasets for subsequent analyses. Please refer to **Table S7** for a comprehensive list of the 113 GWAS datasets of diseases/traits used in our analyses.

### Supplementary Figures

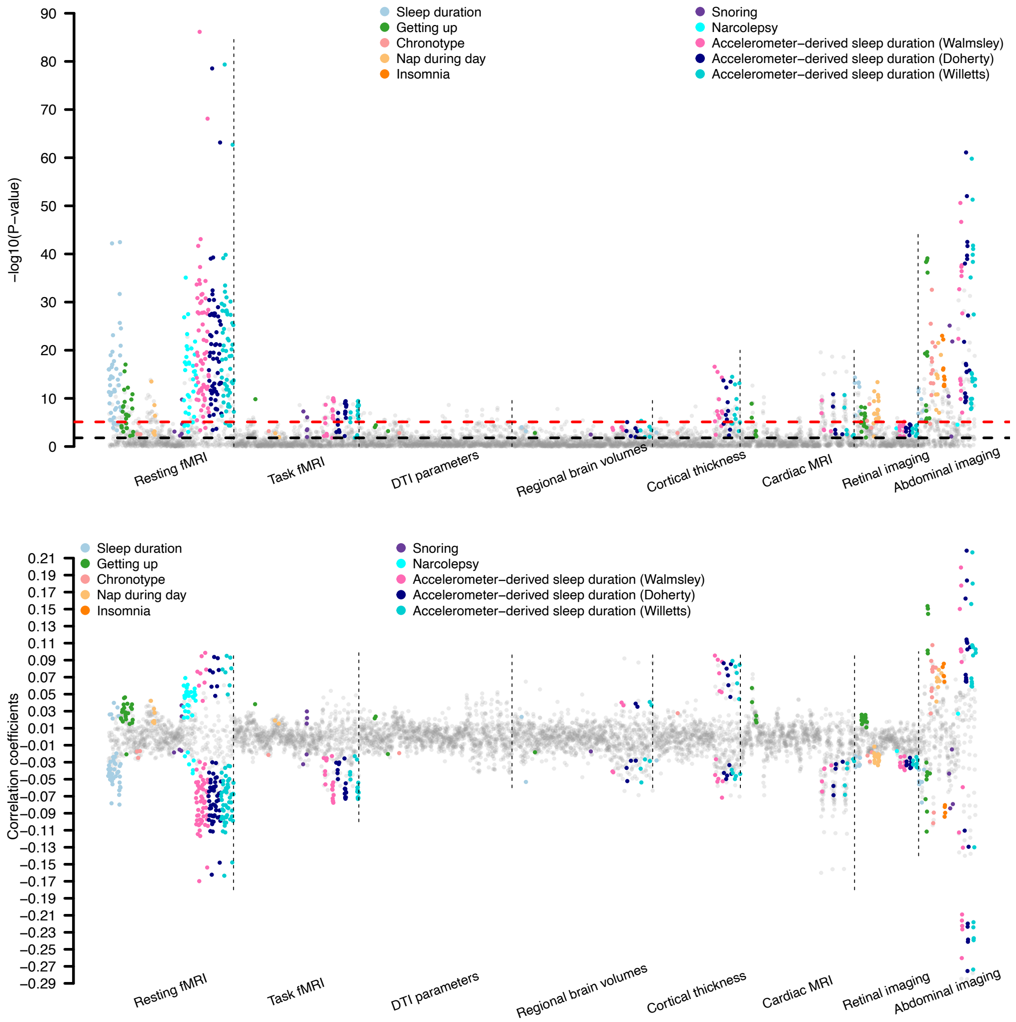


**Fig. S1 Phenotypic associations between sleep and multi-organ imaging traits.**

The -log10(P-value) and correlation coefficients between 10 sleep traits and multi-organ imaging traits, including 101 regional brain volumes, 63 cortical thickness, 110 DTI parameters, 90 resting fMRI traits, 90 task fMRI traits, 82 cardiac MRI traits, 46 derived OCT traits, and 41 abdominal imaging traits. Associations survived the false discovery (FDR) rate of 5% ($P < 1.60 \times 10^{-2}$) and were validated in the two approaches were highlighted with colors. Each sleep trait is labeled with a different color. A red dashed line represents the threshold from Bonferroni correction ($P < 8.03 \times 10^{-6}$), and a black dashed line represents threshold from FDR correction ($P < 1.60\times 10^{-2}$). See Table S1 for more information of these imaging traits.

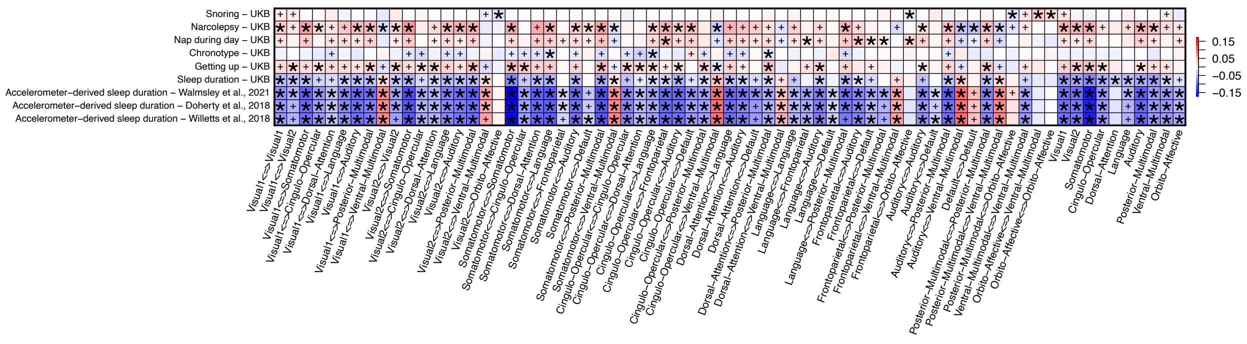


**Fig. S2 Phenotypic associations between sleep and resting fMRI traits.**

We illustrate the correlation coefficients between sleep (*y* axis) and resting fMRI traits (*x* axis). The color represents correlation estimates. The coefficients that pass the false discovery (FDR) rate of 5% ($P < 1.60 \times 10^{-2}$) were marked with asterisk. The coefficients pass the FDR multiple testing but not validated in our approaches were marked with plus sign.

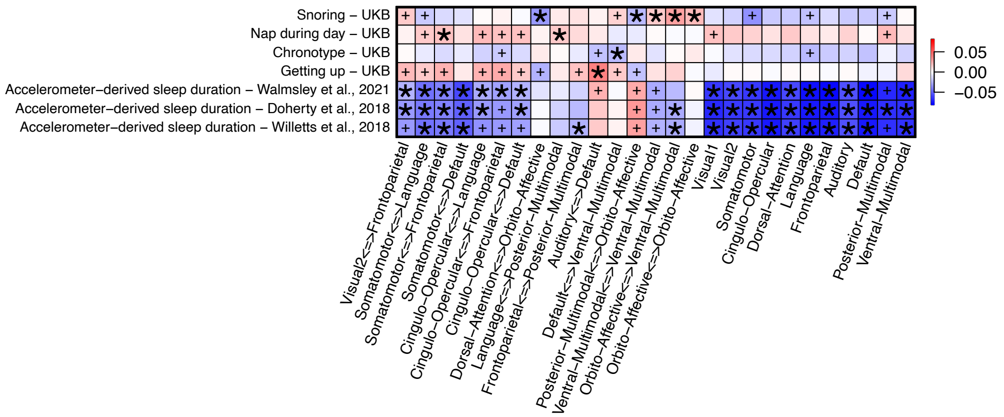


**Fig. S3 Phenotypic associations between sleep and task fMRI traits.**

We illustrate the correlation coefficients between sleep (*y* axis) and task fMRI traits (*x* axis). The color represents correlation estimates. The coefficients that pass the false discovery (FDR) rate of 5% ($P < 1.60 \times 10^{-2}$) were marked with asterisk. The coefficients pass the FDR multiple testing but not validated in our approaches were marked with plus sign.

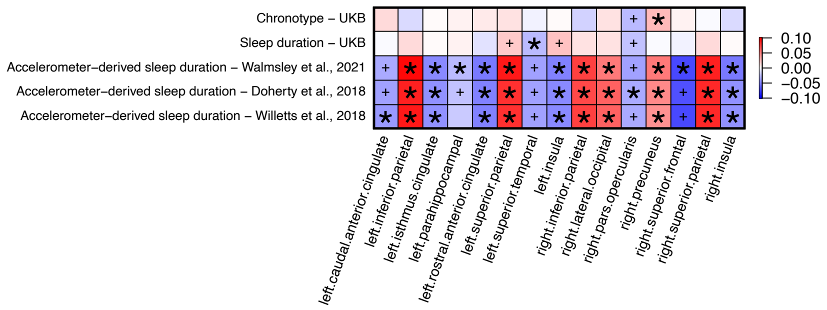


**Fig. S4 Phenotypic associations between sleep and cortical thickness.**

We illustrate the correlation coefficients between sleep (*y* axis) and cortical thickness (*x* axis). The color represents correlation estimates. The coefficients that pass the false discovery (FDR) rate of 5% ($P < 1.60 \times 10^{-2}$) were marked with asterisk. The coefficients pass the FDR multiple testing but not validated in our approaches were marked with plus sign.

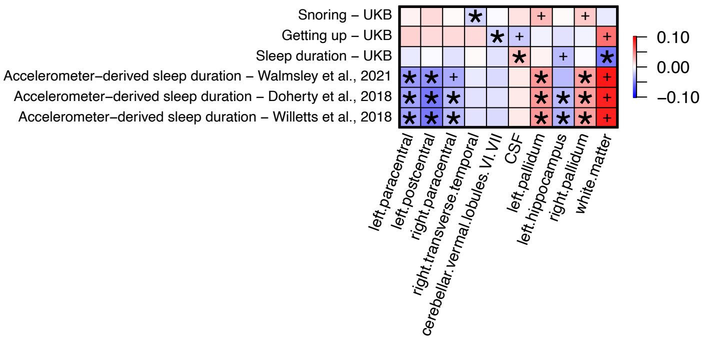


**Fig. S5 Phenotypic associations between sleep and regional brain volumes.**

We illustrate the correlation coefficients between sleep (*y* axis) and regional brain volumes (*x* axis). The color represents correlation estimates. The coefficients that pass the false discovery (FDR) rate of 5% ($P < 1.60 \times 10^{-2}$) were marked with asterisk. The coefficients pass the FDR multiple testing but not validated in our approaches were marked with plus sign.

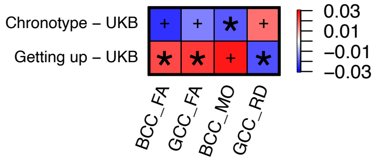


**Fig. S6 Phenotypic associations between sleep and DTI parameters.**

We illustrate the correlation coefficients between sleep (*y* axis) and DTI parameters (*x* axis). The color represents correlation estimates. The coefficients that pass the false discovery (FDR) rate of 5% ($P < 1.60 \times 10^{-2}$) were marked with asterisk. The coefficients pass the FDR multiple testing but not validated in our approaches were marked with plus sign. BCC FA, mean fractional anisotropy (FA) of body of corpus callosum; GCC FA, mean FA of genu of corpus callosum; BCC MD, mean diffusivity (MD) of body of corpus callosum; GCC RD, mean radial diffusivity (RD) of genu of corpus callosum.

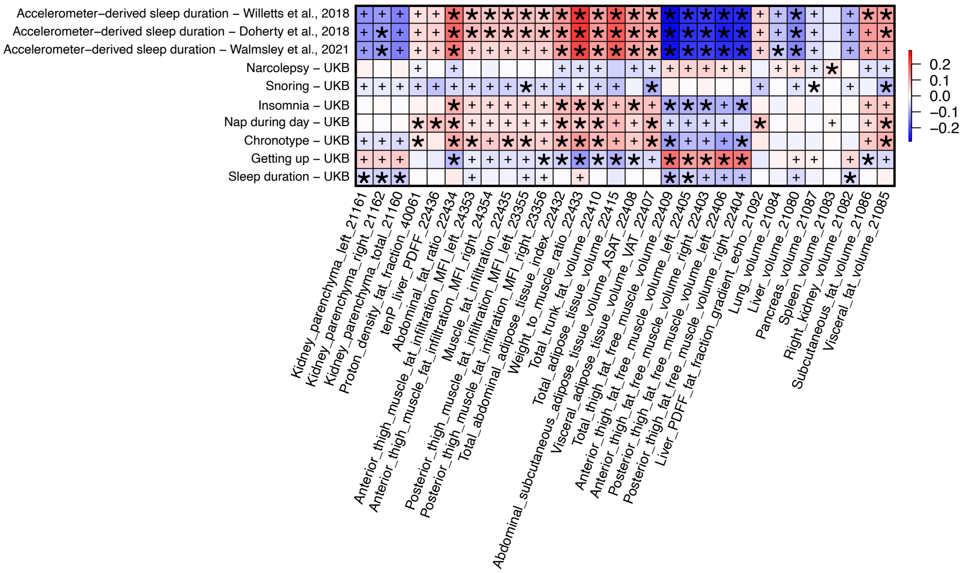


**Fig. S7 Phenotypic associations between sleep and abdominal MRI traits.**

We illustrate the correlation coefficients between sleep (*y* axis) and abdominal MRI traits (*x* axis). The color represents correlation estimates. The coefficients that pass the false discovery (FDR) rate of 5% ($P < 1.60 \times 10^{-2}$) were marked with asterisk. The coefficients pass the FDR multiple testing but not validated in our approaches were marked with plus sign.

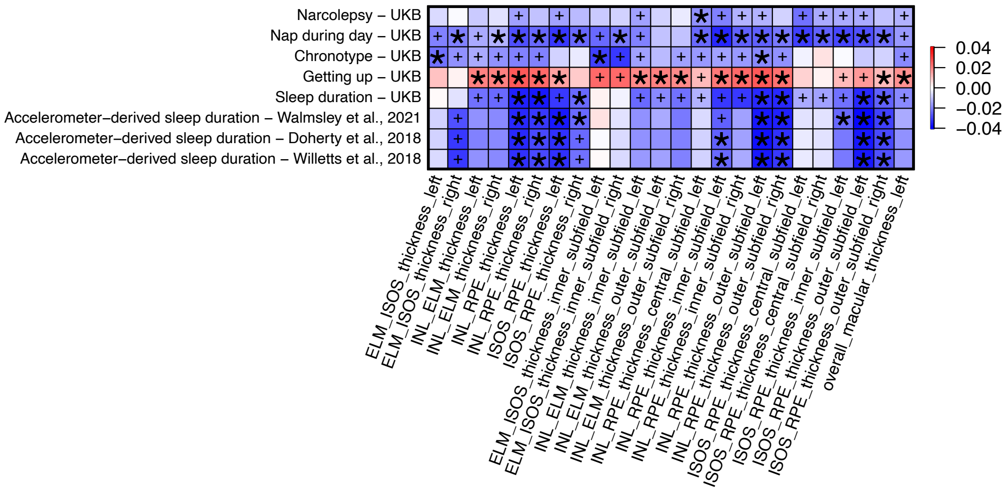


**Fig. S8 Phenotypic associations between sleep and retinal imaging traits.**

We illustrate the correlation coefficients between sleep (*y* axis) and eye imaging traits (*x* axis). The color represents correlation estimates. The coefficients that pass the false discovery (FDR) rate of 5% ($P < 1.60 \times 10^{-2}$) were marked with asterisk. The coefficients pass the FDR multiple testing but not validated in our approaches were marked with plus sign.

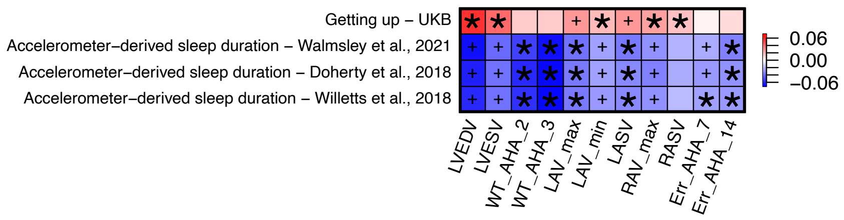


**Fig. S9 Phenotypic associations between sleep and cardiac MRI traits.**

We illustrate the correlation coefficients between sleep (*y* axis) and cardiac MRI traits (*x* axis). The color represents correlation estimates. The coefficients that pass the false discovery (FDR) rate of 5% ($P < 1.60 \times 10^{-2}$) were marked with asterisk. The coefficients pass the FDR multiple testing but not validated in our approaches were marked with plus sign.

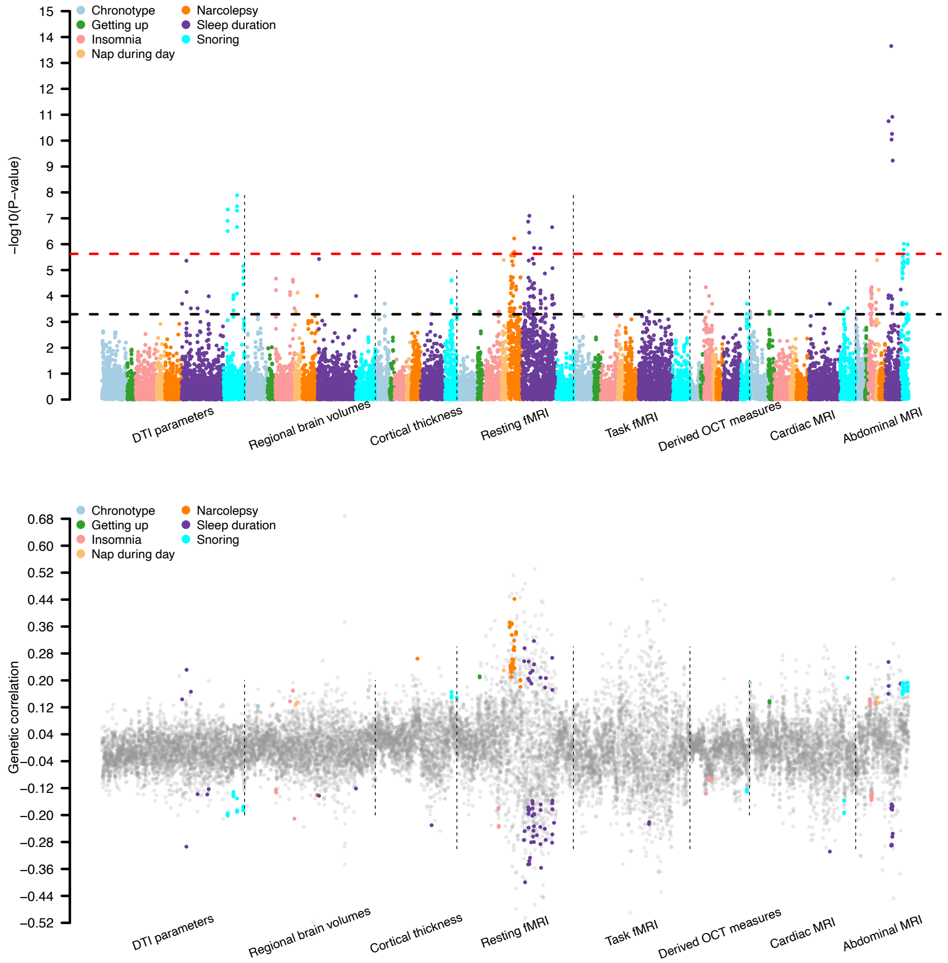


**Fig. S10 Genetic correlations between sleep and multi-organ imaging traits.**

The -log10(P-value) and genetic correlation between GWAS of 34 sleep traits and multi-organ imaging traits, including 101 regional brain volumes, 63 cortical thickness, 110 DTI parameters, 90 resting fMRI traits, 90 task fMRI traits, 82 cardiac MRI traits, 46 derived OCT traits, and 41 abdominal MRI traits. Each sleep trait is labeled with a different color. A red dashed line represents the threshold from Bonferroni correction ($P < 2.36\times 10^{-6}$), and a black dashed line represents threshold from FDR correction ($P < 5.08\times 10^{-4}$). Associations survived the false discovery (FDR) rate of 5% ($P < 5.08\times 10^{-4}$) were highlighted. See Table S1 and Table S3 for more information on GWAS of imaging traits and sleep traits, respectively.

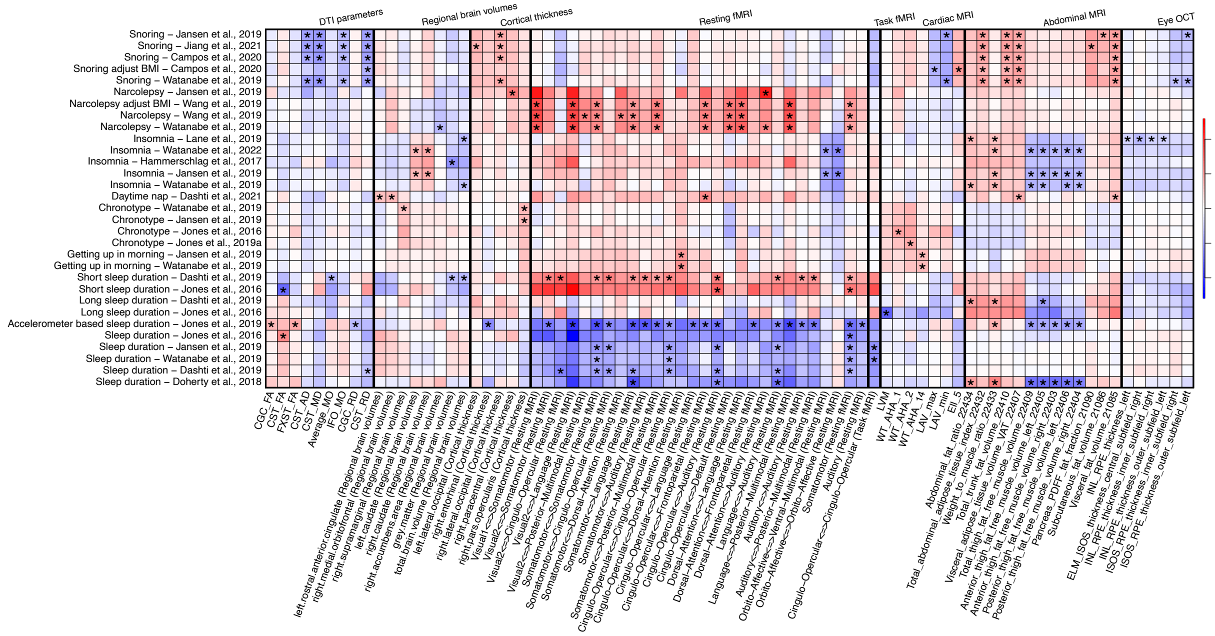


**Fig. S11 Genetic correlations between sleep and multi-organ imaging traits.**

We illustrate the genetic correlation between GWAS of 34 sleep traits (*y* axis) and 623 multi-organ imaging traits (*x* axis), including 101 regional brain volumes, 63 cortical thickness, 110 DTI parameters, 90 resting fMRI traits, 90 task fMRI traits, 82 cardiac MRI traits, 46 derived OCT traits, and 41 abdominal MRI traits. The color represents correlation estimates. The coefficients that pass the false discovery (FDR) rate of 5% ($P < 5.08 \times 10^{-4}$) were marked with asterisk.

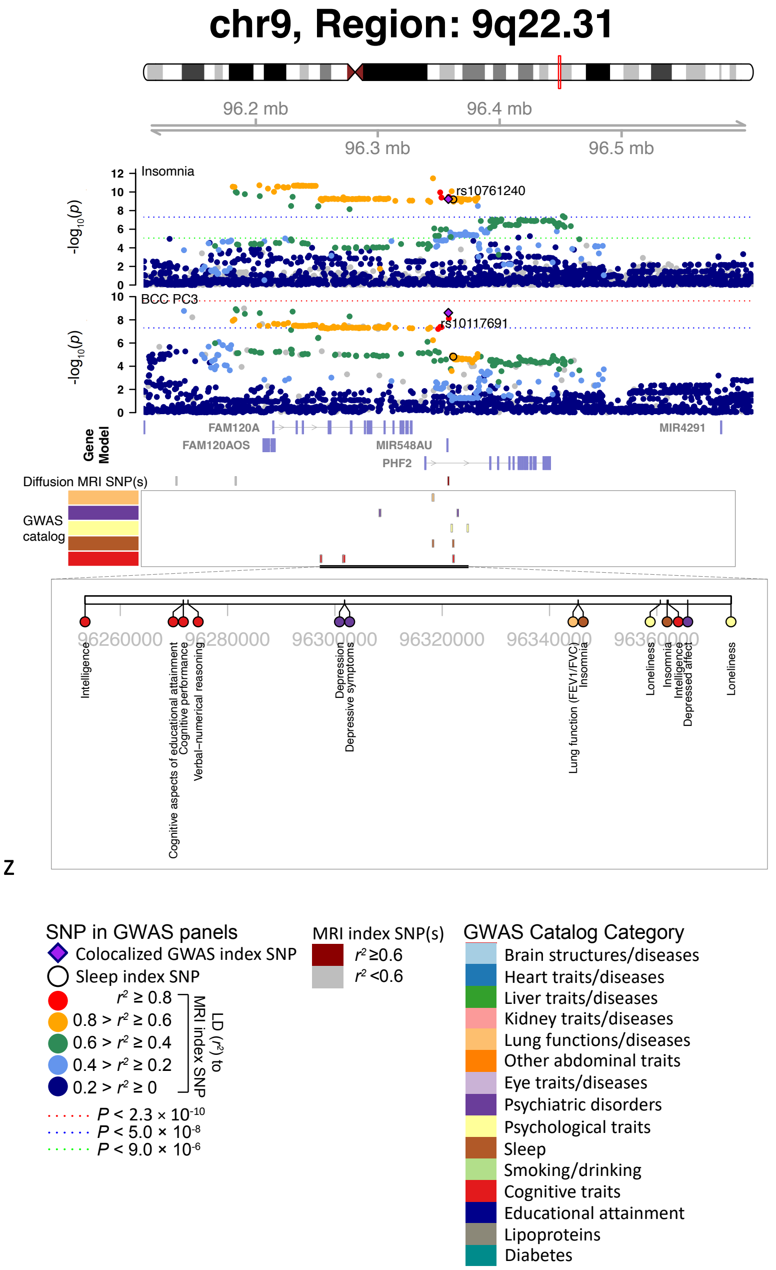


**Fig. S12 Selected genetic locus that was associated with both insomnia and DTI parameters.**

In 9q22.31, we observed the shared association (LD $r^2 \geq 0.6$) between insomnia (index variant rs10761240) and BCC PC3 (index variant rs10117691). We also observed the shared association with lung function, depression, and intelligence. BCC PC3, the third PC of FA in body of corpus callosum in brain diffusion MRI. The posterior probability of Bayesian colocalization analysis for the shared causal variant hypothesis (PPH4) is 85.4%.

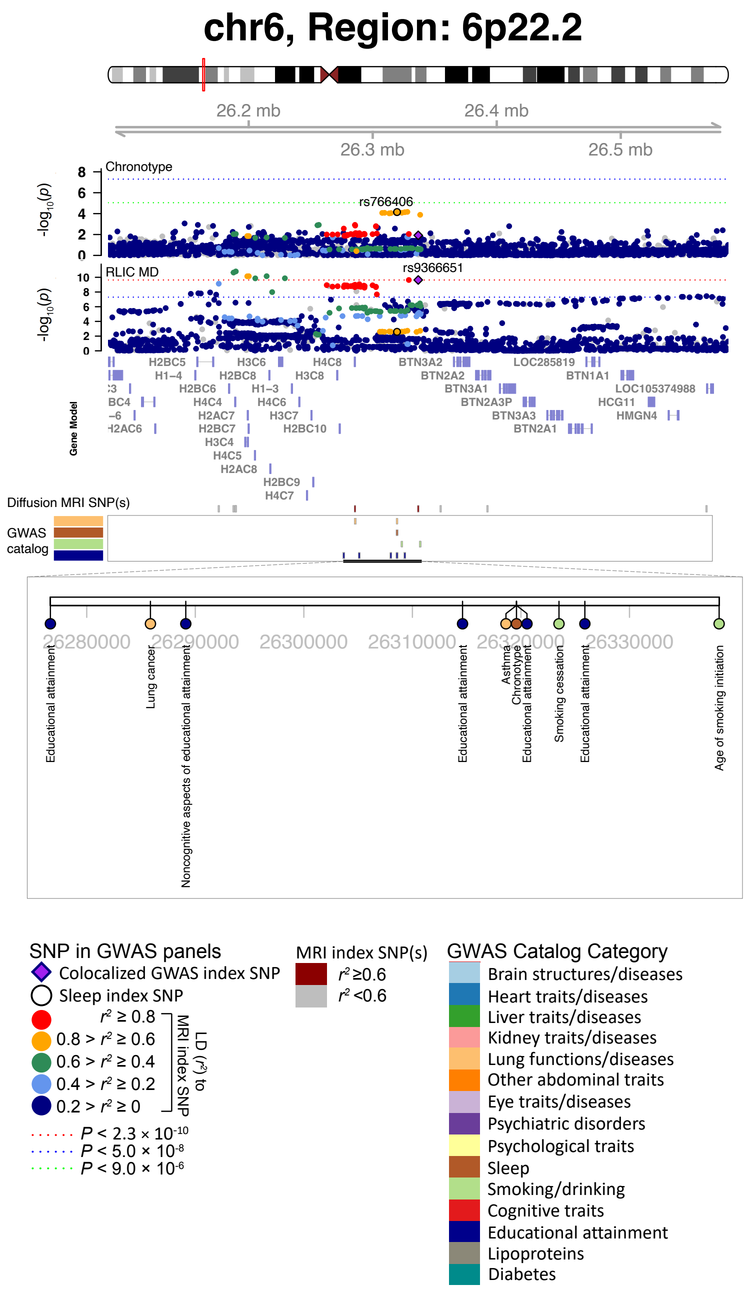


**Fig. S13 Selected genetic locus that was associated with both chronotype and DTI parameters.**

In 6p22.2, we observed the shared association (LD $r^2 \geq 0.6$) between chronotype (index variant rs766406) and RLIC MD (index variant rs9366651). We also observed the shared association with asthma, lung cancer, age of smoking initiation, and educational attainment. RLIC MD, mean MD of retrolenticular part of internal capsule in brain diffusion MRI.

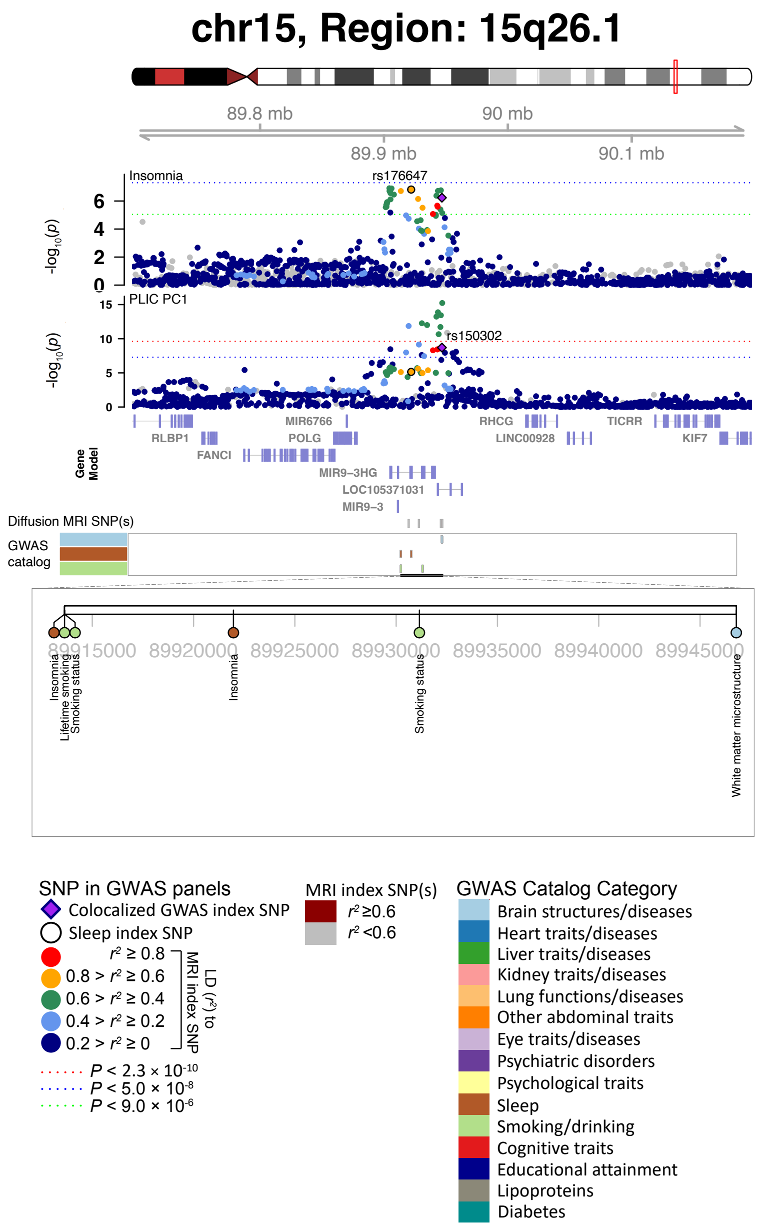


**Fig. S14 Selected genetic locus that was associated with both insomnia and DTI parameters.**

In 15q26.1, we observed the shared association (LD $r^2 \geq 0.6$) between insomnia (index variant rs176647) and PLIC PC1 (index variant rs150302). We also observed the shared association with smoking status. PLIC PC1, the first PC of FA in posterior limb of internal capsule in brain diffusion MRI. The posterior probability of Bayesian colocalization analysis for the shared causal variant hypothesis (PPH4) is 87.1%.

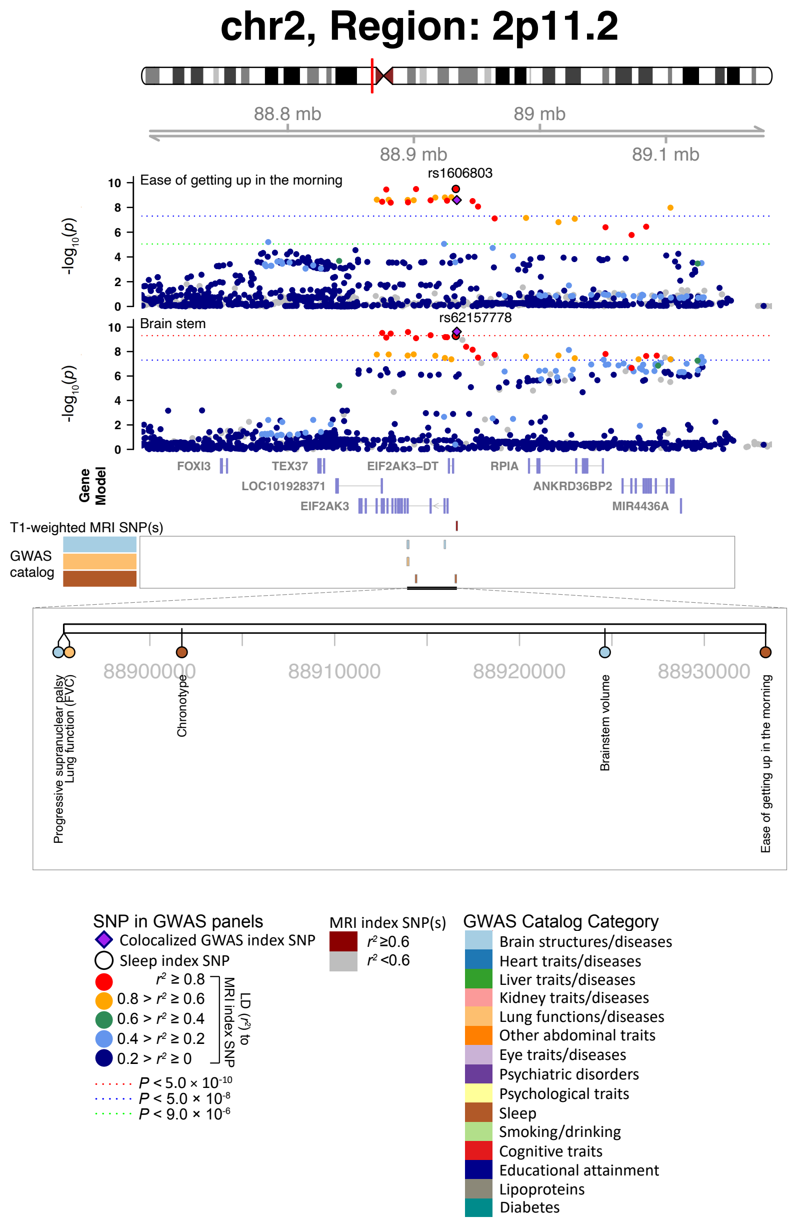


**Fig. S15 Selected genetic loci that were associated with both ease of getting up in the morning and regional brain volumes.**

In 2p11.2, we observed the shared association (LD $r^2 \geq 0.6$) between ease of getting up in the morning (index variant rs1606803) and volume of brain stem (index variant rs62157778). The posterior probability of Bayesian colocalization analysis for the shared causal variant hypothesis (PPH4) is 0.983.

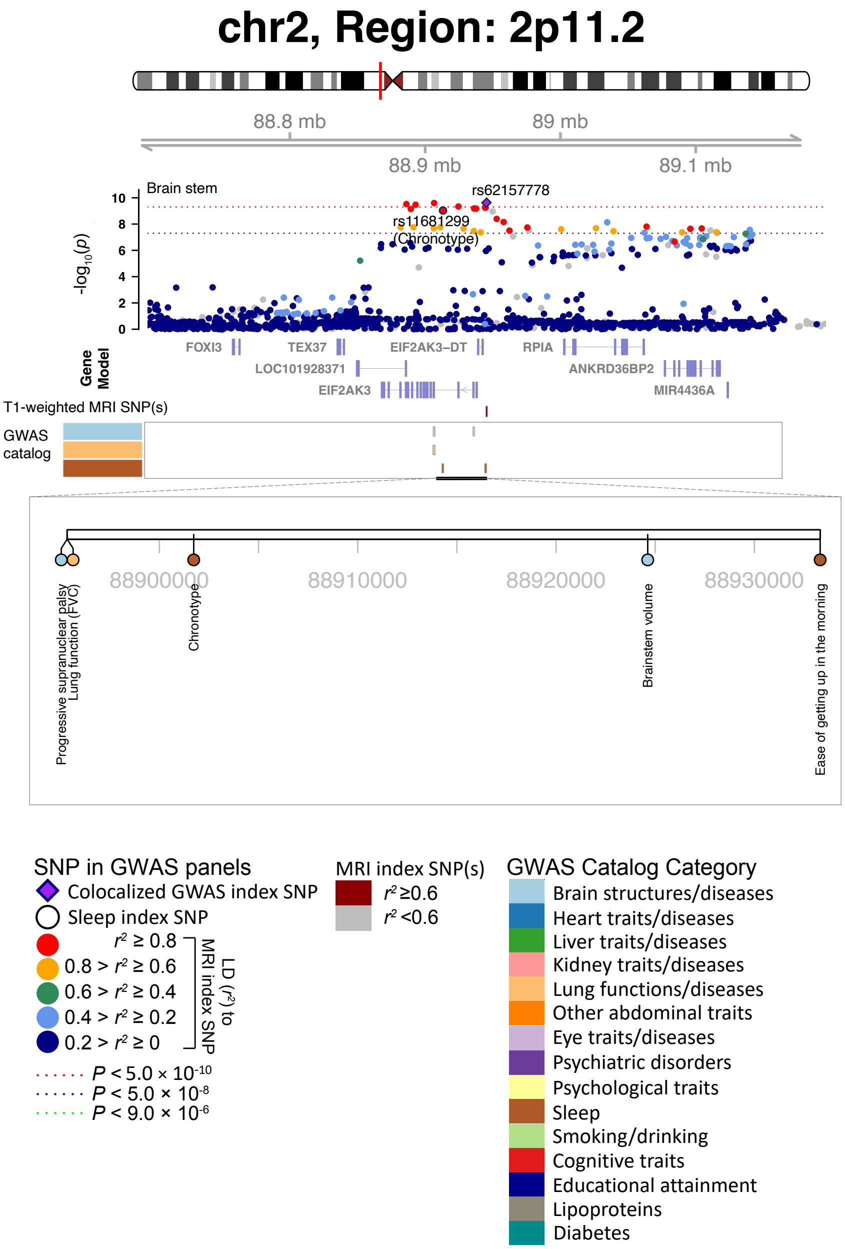


**Fig. S16 Selected genetic loci that were associated with both chronotype and regional brain volumes.**

In 2p11.2, we observed the shared association (LD $r^2 \geq 0.6$) between chronotype (index variant rs11681299) and volume of brain stem (index variant rs62157778).

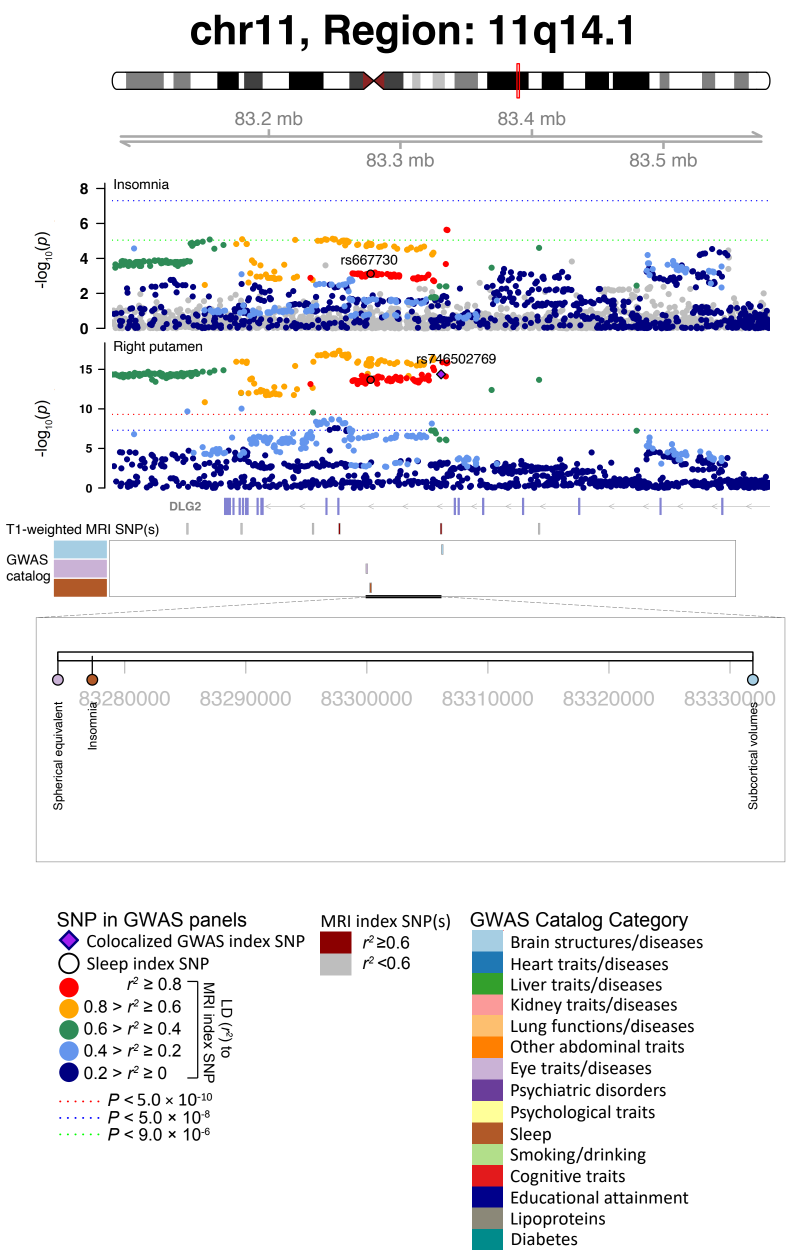


**Fig. S17 Selected genetic loci that were associated with both chronotype and regional brain volumes.**

In 11q14.1, we observed the shared association (LD $r^2 \geq 0.6$) between insomnia (index variant rs667730) and volume of the left putamen (index variant rs746502769). The posterior probability of Bayesian colocalization analysis for the shared causal variant hypothesis (PPH4) is 0.9061. We also observed the shared associations with neuroticism, depressive symptoms, insomnia, and ease of getting up in the morning.

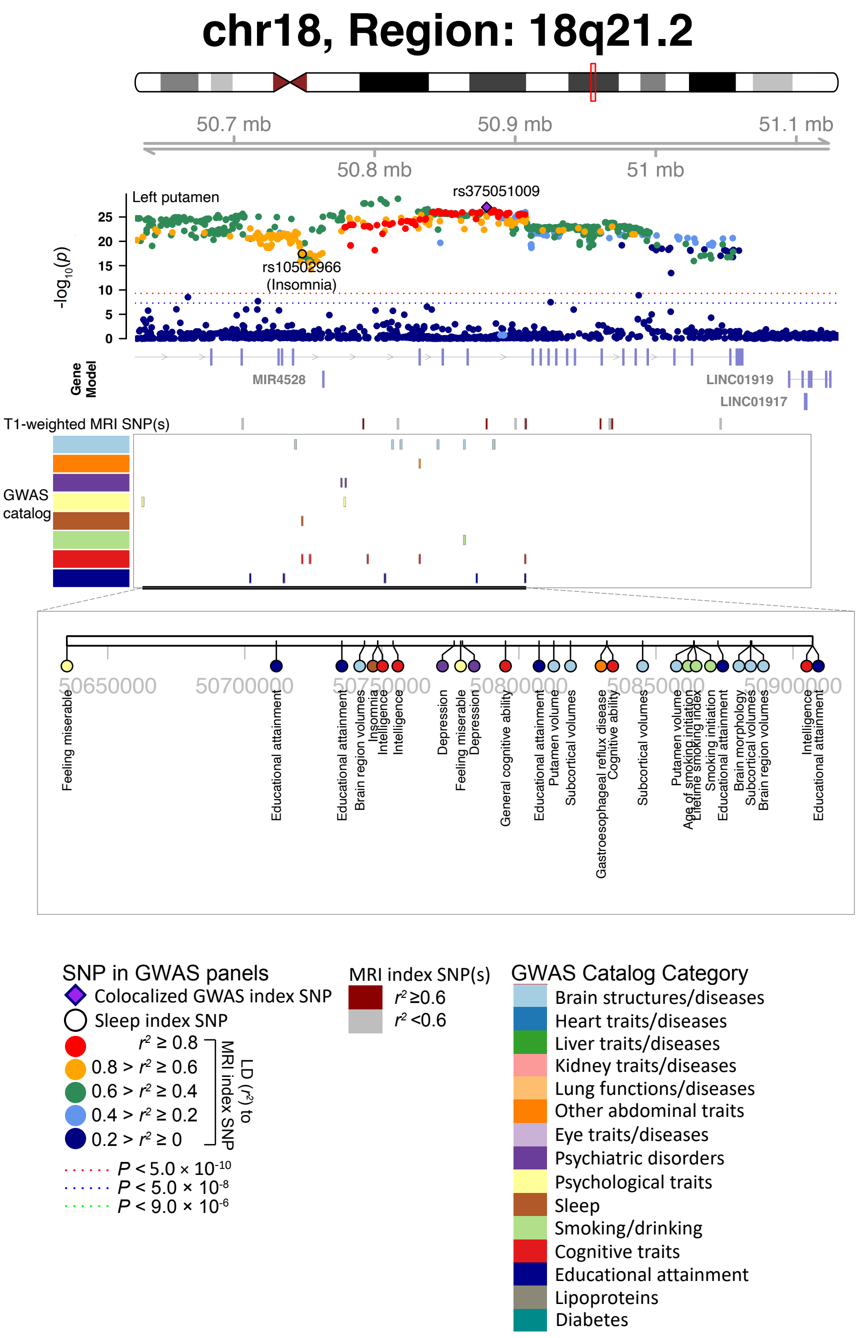


**Fig. S18 Selected genetic loci that were associated with both insomnia and regional brain volumes.**

In 18q21.2, we observed the shared association (LD $r^2 \geq 0.6$) between insomnia (index variant rs10502966) and volume of the left putamen (index variant rs375051009). We also observed the shared associations with educational attainment, intelligence, depression, cognitive ability, smoking initiation, and gastroesophageal reflux disease.

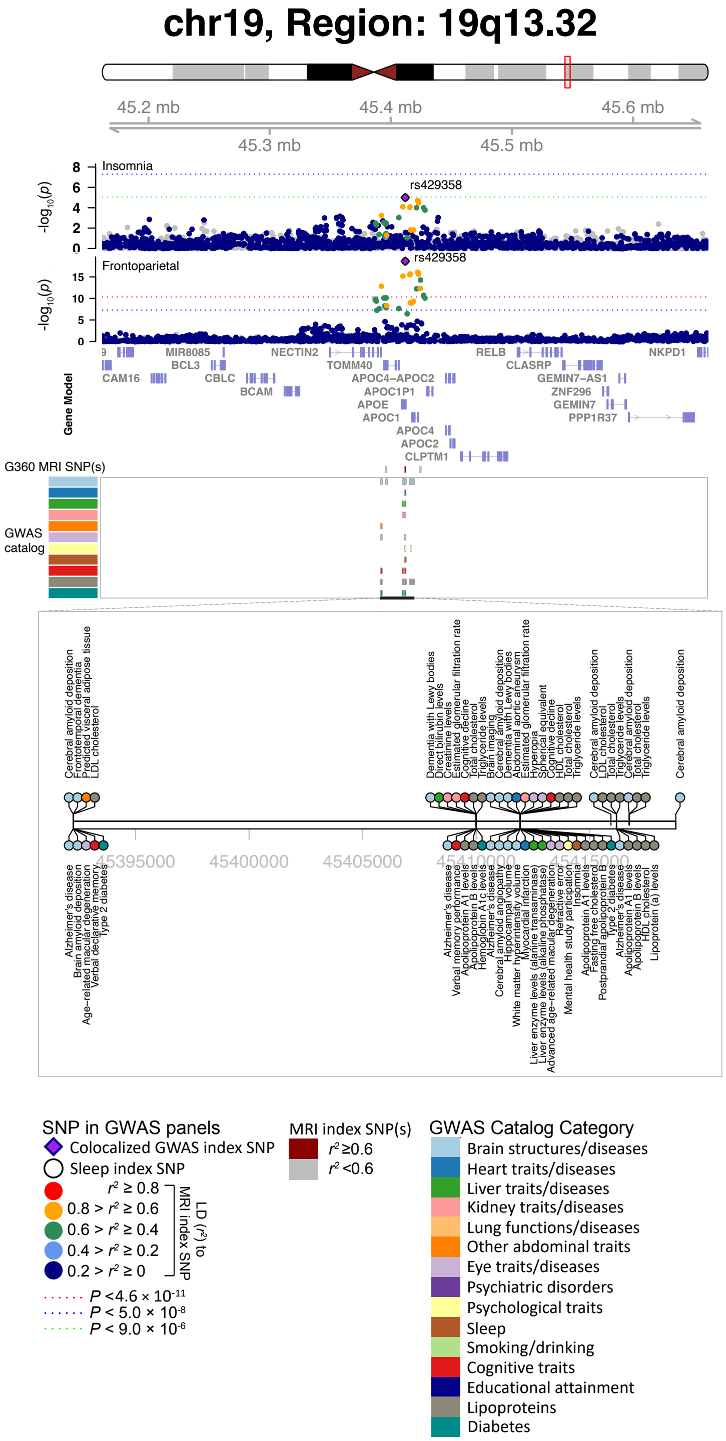


**Fig. S19 Selected genetic loci that were associated with both insomnia and resting fMRI traits.**

In 19q13.32, we observed the shared association (LD $r^2 \geq 0.6$) between insomnia (index variant rs429358) and resting functional connectivity in the frontoparietal network (index variant rs429358). The posterior probability of Bayesian colocalization analysis for the shared causal variant hypothesis (PPH4) is 0.980. We also observed the shared associations with type 2 diabetes, LDL cholesterol, cognitive decline, age-related macular-degeneration, and Alzheimer's disease.

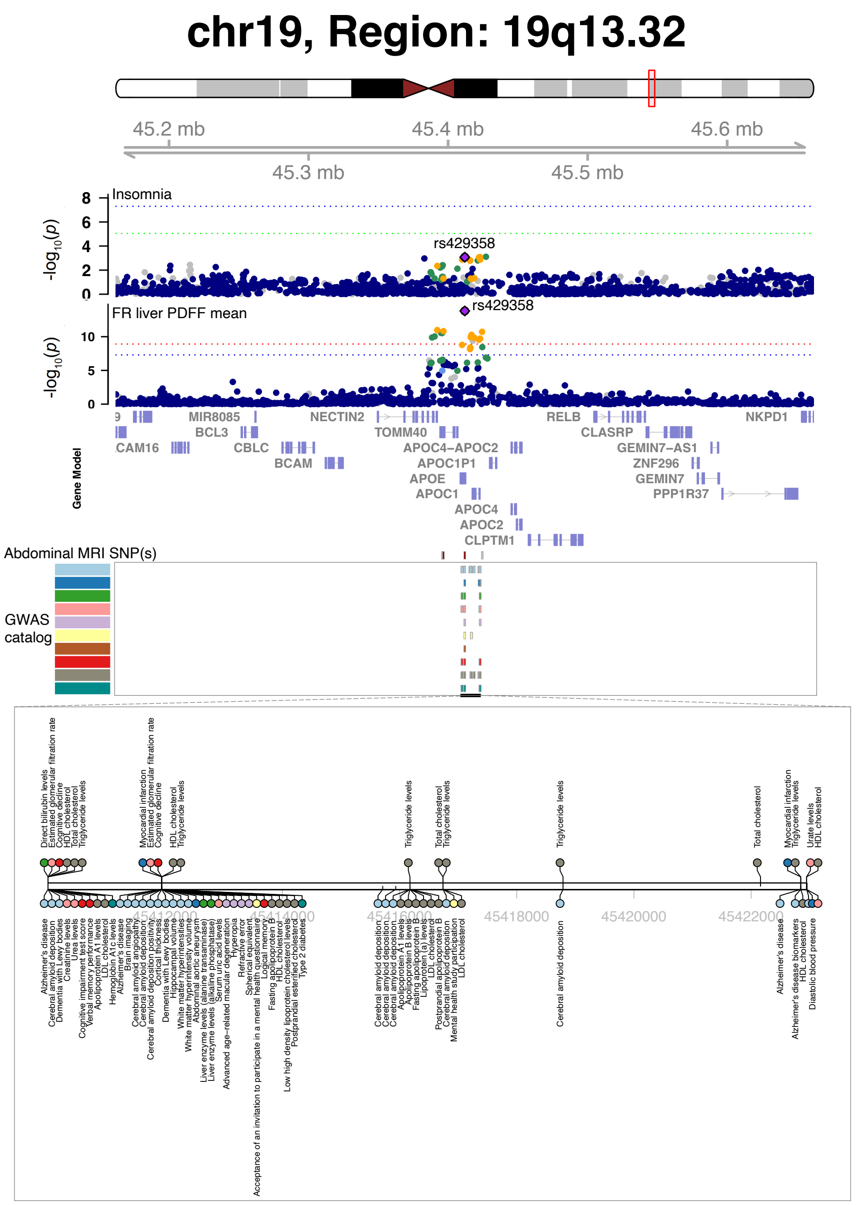


**Fig. S20 Selected genetic loci that were associated with both insomnia and abdominal MRI traits.**

In 19q13.32, we observed the shared association (LD $r^2 \geq 0.6$) between insomnia (index variant rs429358) and the average proton density fat fraction (PDFF) in the liver (index variant rs429358). The posterior probability of Bayesian colocalization analysis for the shared causal variant hypothesis (PPH4) is 0.980. We also observed the shared associations with type 2 diabetes, LDL cholesterol, cognitive decline, age-related macular-degeneration, and Alzheimer's disease.

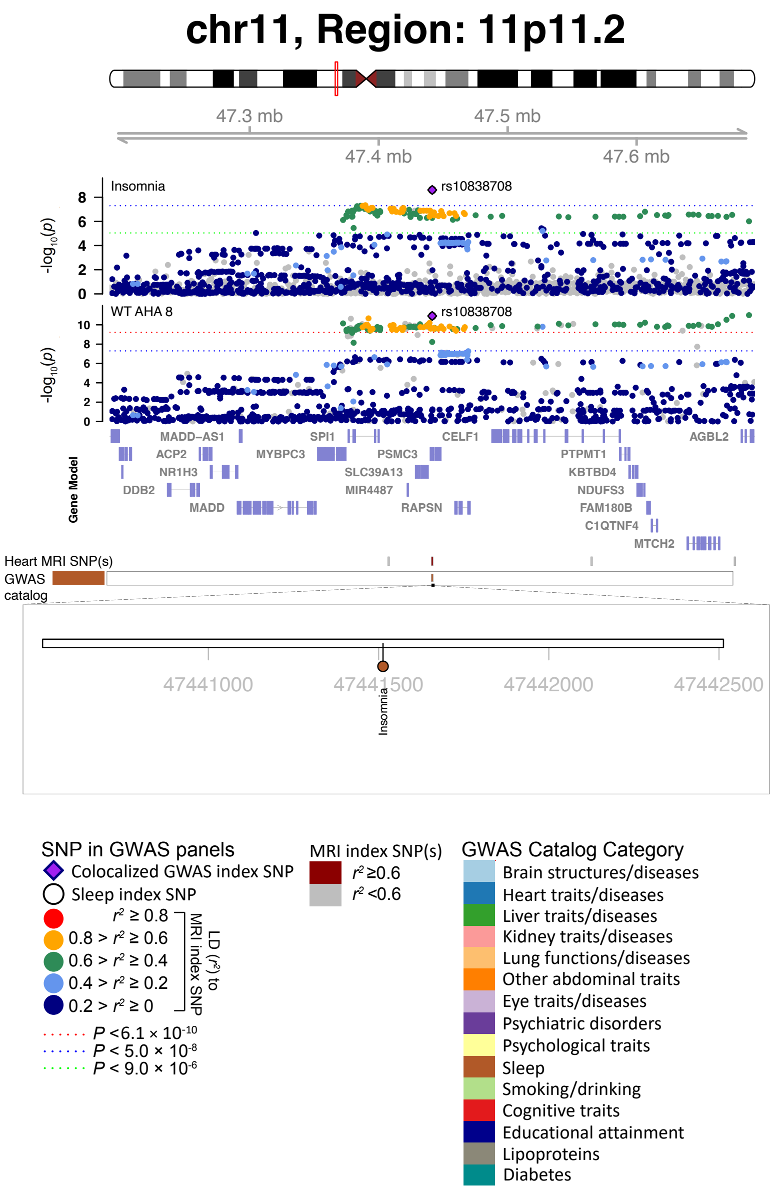


**Fig. S21 Selected genetic loci that were associated with both insomnia and cardiac MRI traits.**

In 11p11.2, we observed the shared association (LD $r^2 \geq 0.6$) between insomnia (index variant rs10838708) and WT AHA 8 (index variant rs10838708). WT AHA 8, regional myocardial-wall thickness at end-diastole.

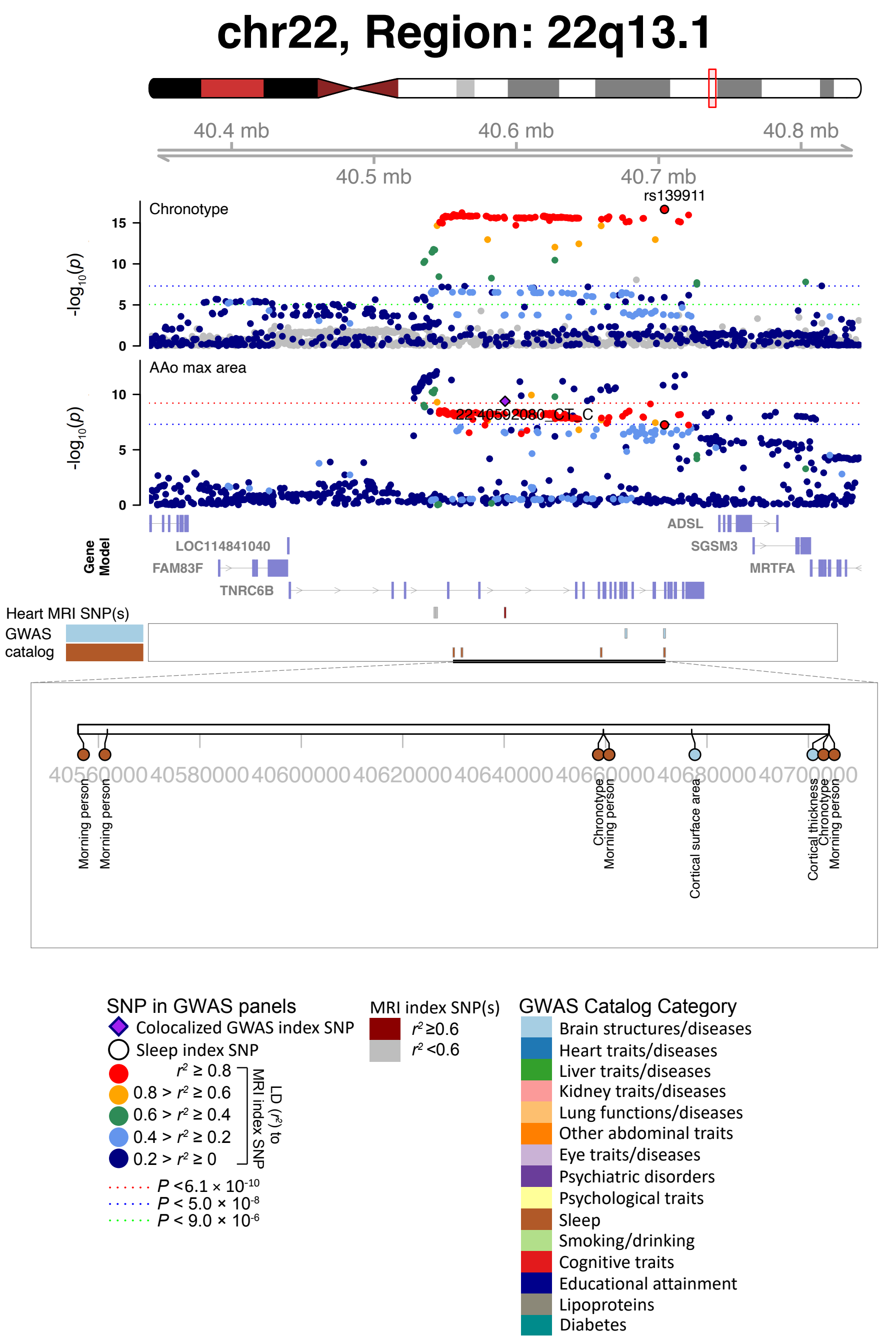


**Fig. S22 Selected genetic loci that were associated with both chronotype and cardiac MRI traits.**

In 22q13.1, we observed the shared association (LD $r^2 \geq 0.6$) between chronotype (index variant rs139911) and AAo max area (index variant 22:40592080_CT_C). AAo max area, ascending aorta maximum area.

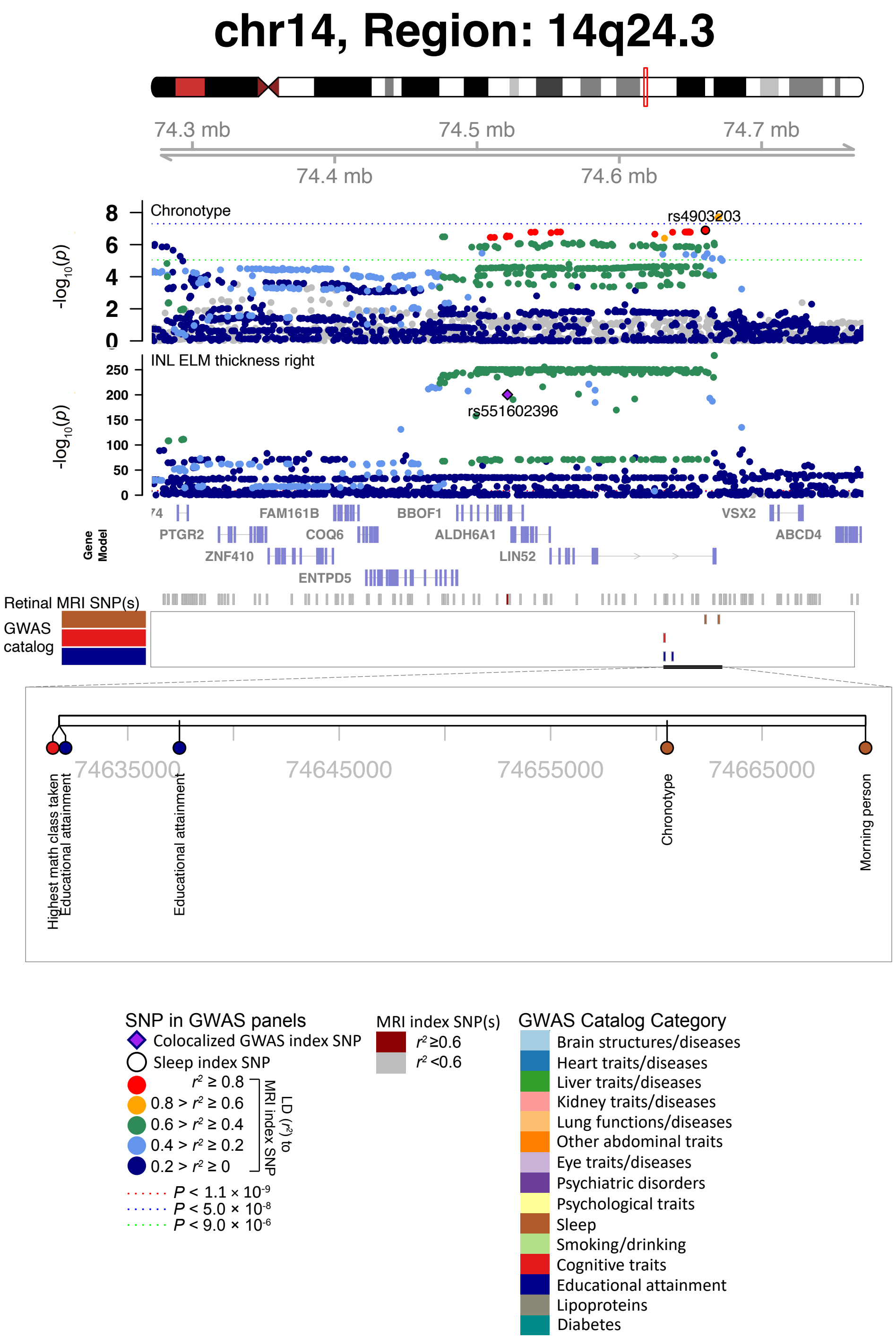


**Fig. S23 Selected genetic loci that were associated with both chronotype and derived OCT measures.**

In 14q24.3, we observed the shared association (LD $r^2 \geq 0.6$) between chronotype (index variant rs4903203) and INL ELM thickness right (index variant rs551602396). We also observed the shared association with highest math class taken and educational attainment.

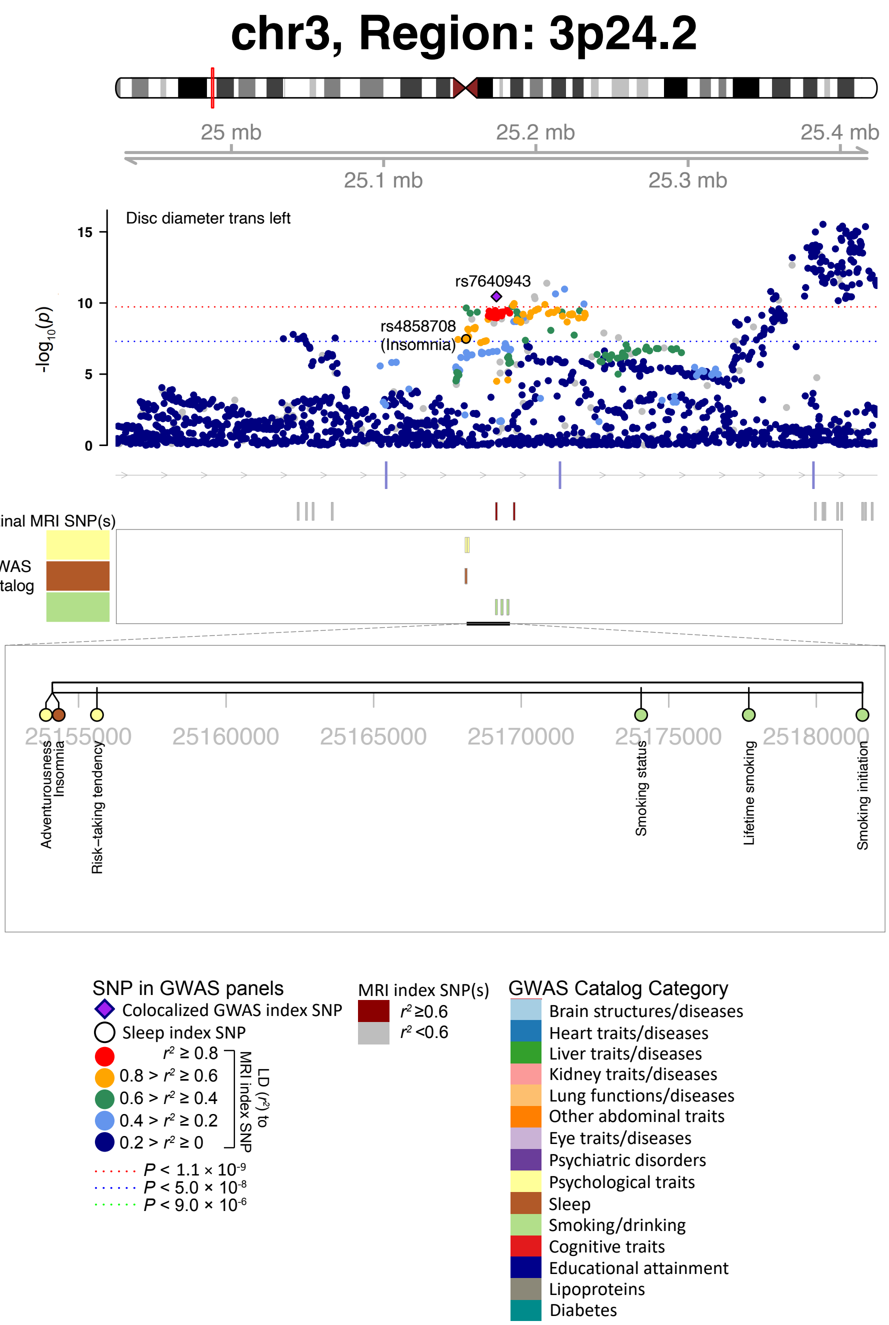


**Fig. S24 Selected genetic loci that were associated with both insomnia and derived OCT measures.**

In 3p24.2, we observed the shared association (LD $r^2 \geq 0.6$) between insomnia (index variant rs4858708) and disc diameter trans left (index variant rs7640943). We also observed the shared association with risk-taking tendency and smoking status.

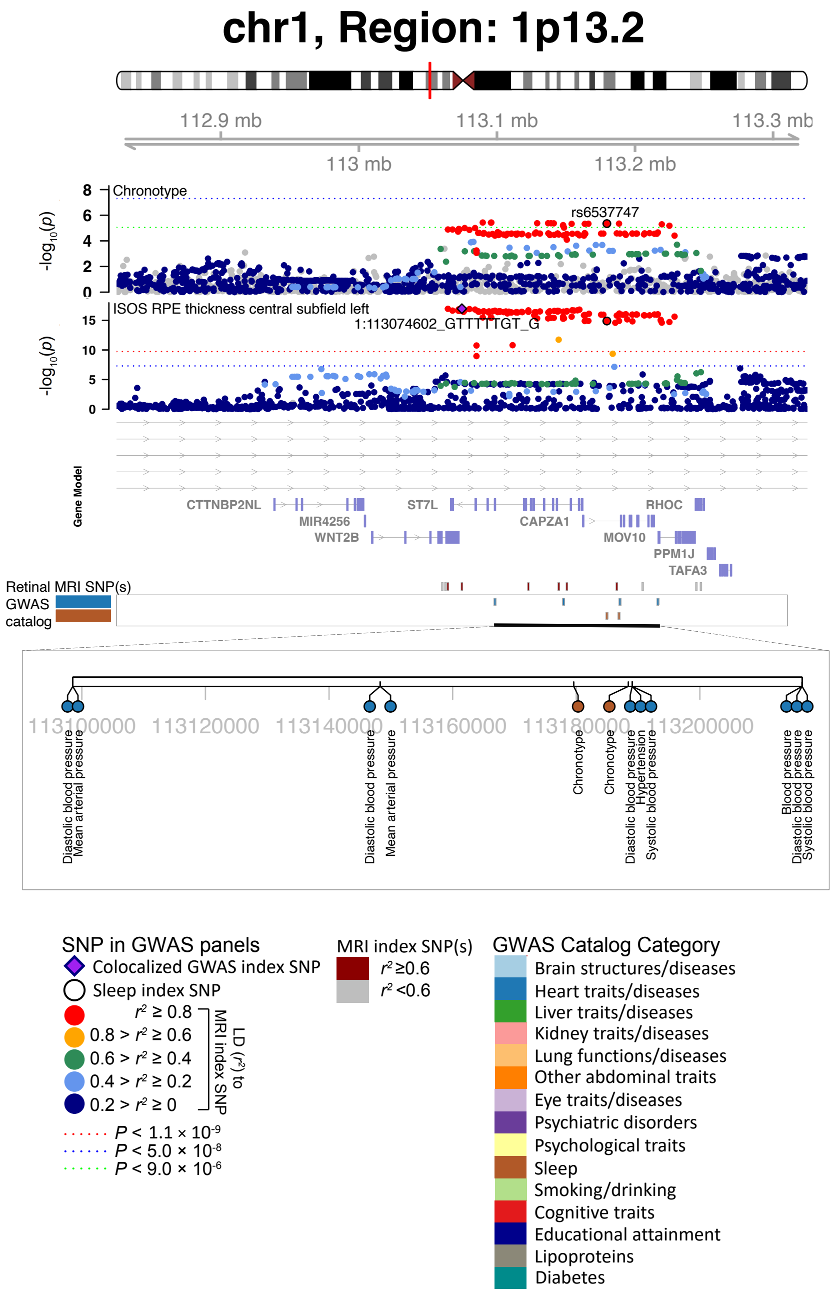


**Fig. S25 Selected genetic loci that were associated with both chronotype and derived OCT measures.**

In 1p13.2, we observed the shared association (LD $r^2 \geq 0.6$) between chronotype (index variant rs6537747) and ISOS RPE thickness central subfield left (index variant 1:113074602_GTTTTTGT_G). We also observed the shared association with mean arterial pressure, blood pressure, and hypertension.

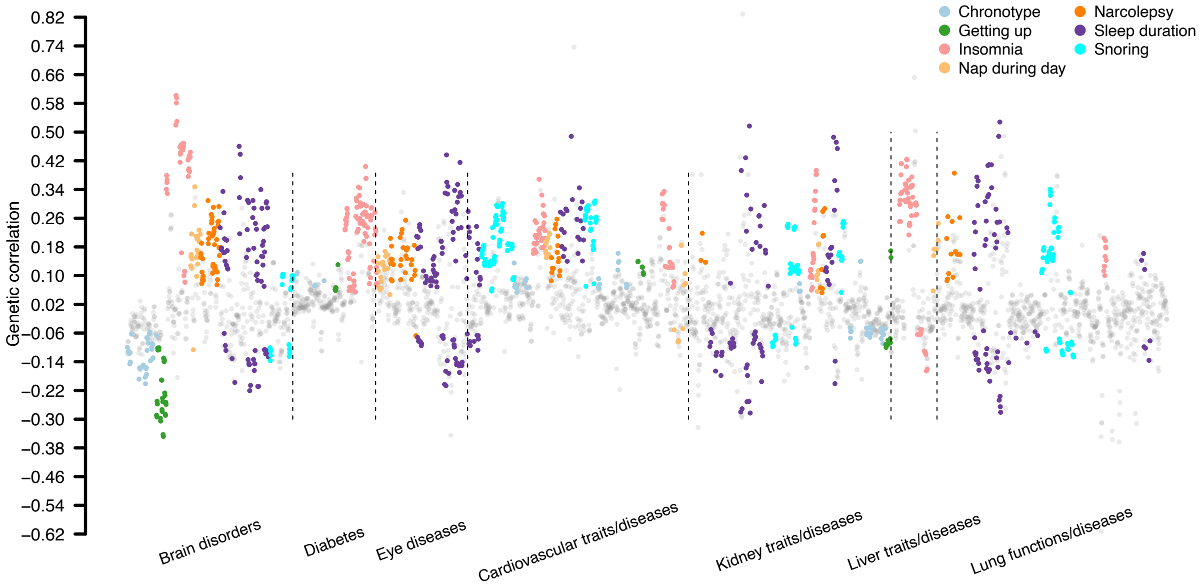


**Fig. S26 Genetic correlations between sleep and diseases/traits.**

The correlation coefficients between GWAS of 34 sleep traits and 7 groups of diseases/traits, including seven brain disorders, 13 cardiovascular diseases/traits, two diabetes, 2 eye diseases, ten kidney traits/diseases, four liver traits/diseases, and 11 lung functions/diseases. Each trait is labeled with a different color. Correlations survived the false discovery (FDR) rate of 5% ($P < 1.55 \times 10^{-2}$) were highlighted. See Table S7 and Table S3 for more information on GWAS of multi-organ traits/diseases and sleep traits, respectively.

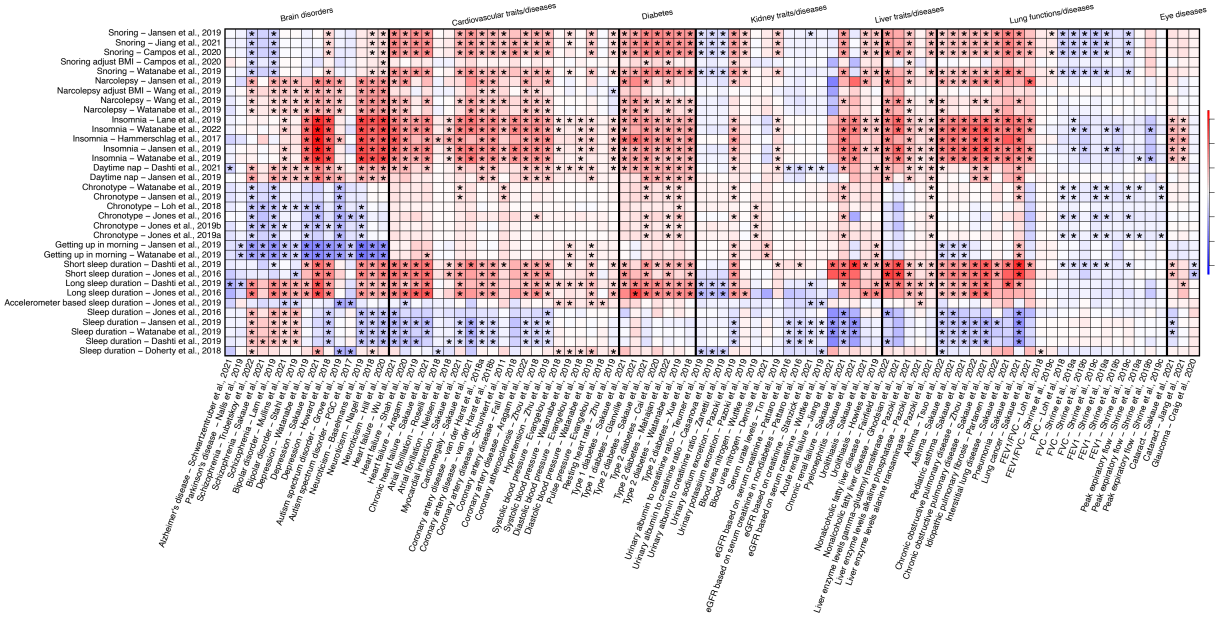


**Fig. S27 Genetic correlations between sleep and diseases/traits.**

We illustrate the genetic correlations between sleep (*y* axis) and diseases/traits (*x* axis). The color represents correlation estimates. The coefficients that pass the false discovery (FDR) rate of 5% ($P < 1.55 \times 10^{-2}$) were marked with asterisk.

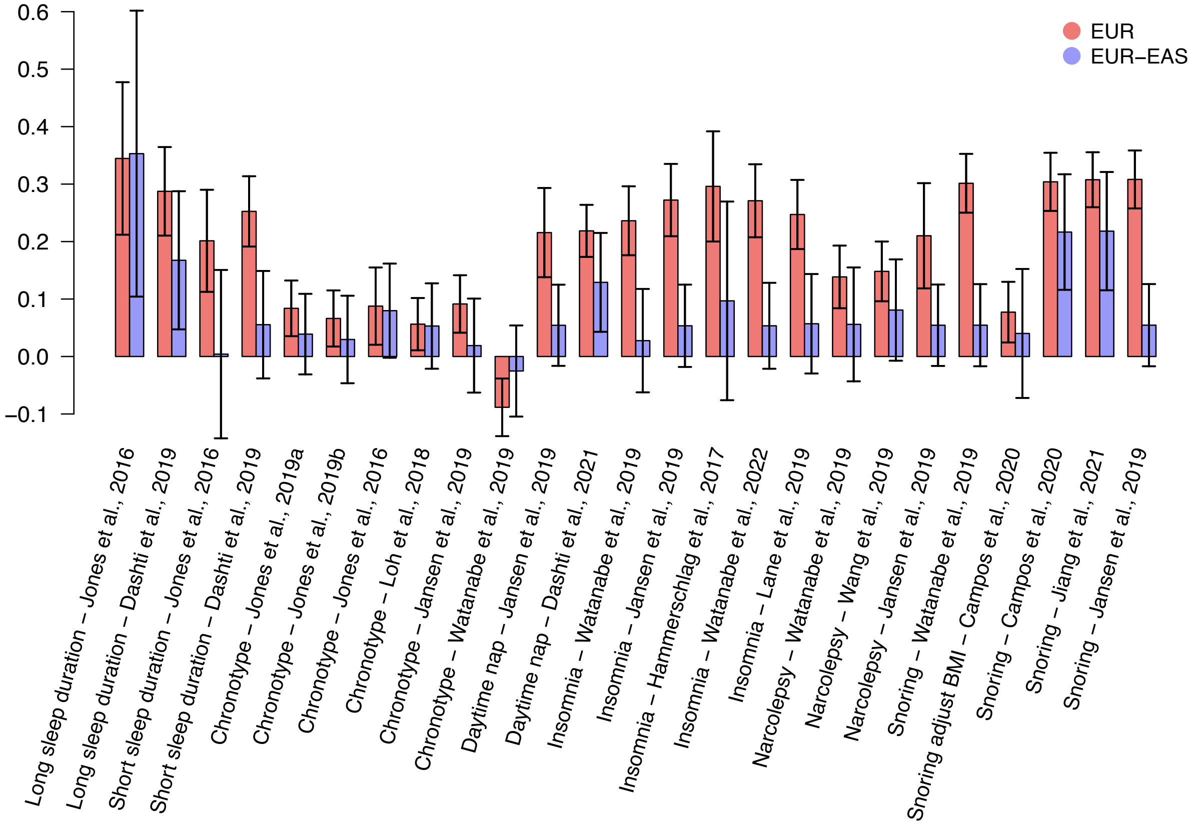


**Fig. S28 Cross-ancestry genetic correlations between sleep and type 2 diabetes.**

We illustrate the cross-ancestry genetic correlation between sleep and type 2 diabetes. The red bars represent genetic correlations between sleep and type 2 diabetes within the European population, while the blue bars represent genetic correlations between sleep (European) and type 2 diabetes (East Asian). Sleep traits whose genetic correlations pass the false discovery (FDR) rate of 5% ($P < 1.81 \times 10^{-2}$) with type 2 diabetes within the European population were plotted.

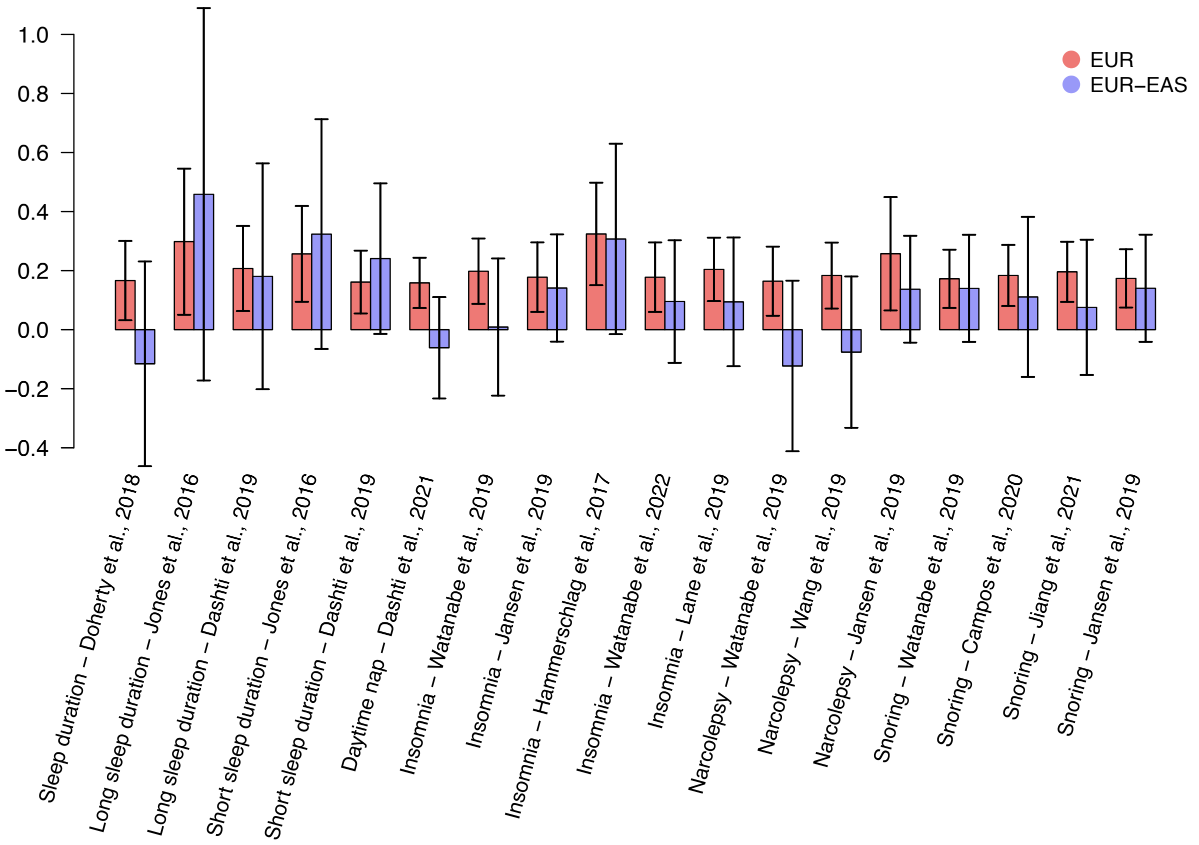


**Fig. S29 Cross-ancestry genetic correlations between sleep and type 1 diabetes.**
We illustrate the cross-ancestry genetic correlation between sleep and type 1 diabetes. The red bars represent genetic correlations between sleep and type 1 diabetes within the European population, while the blue bars represent genetic correlations between sleep (European) and type 1 diabetes (East Asian). Sleep traits whose genetic correlations pass the false discovery (FDR) rate of 5% ($P < 1.81 \times 10^{-2}$) with type 1 diabetes within the European population were plotted.

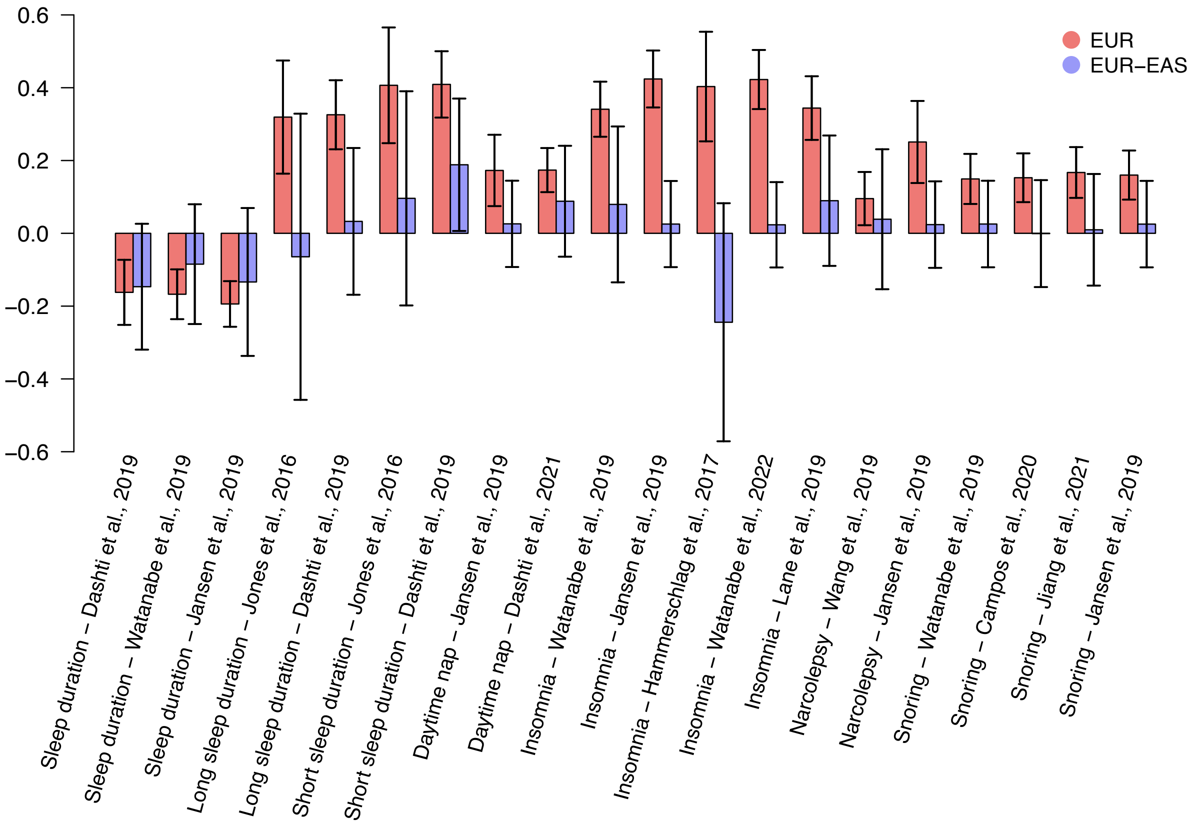


**Fig. S30 Cross-ancestry genetic correlations between sleep and chronic obstructive pulmonary disease.**

We illustrate the cross-ancestry genetic correlation between sleep and chronic obstructive pulmonary disease (COPD). The red bars represent genetic correlations between sleep and COPD within the European population, while the blue bars represent genetic correlations between sleep (European) and COPD (East Asian). Sleep traits whose genetic correlations pass the false discovery (FDR) rate of 5% ($P < 1.81 \times 10^{-2}$) with COPD within the European population were plotted.

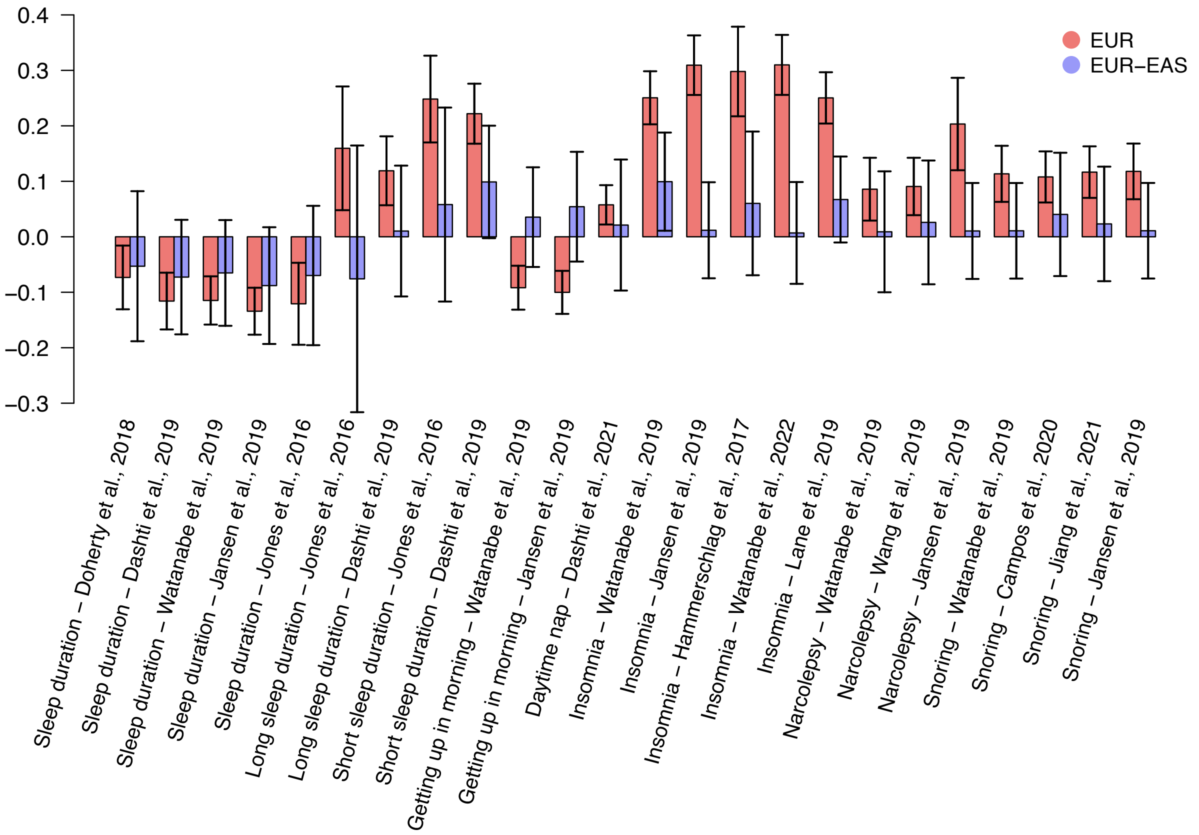


**Fig. S31 Cross-ancestry genetic correlations between sleep and asthma disease.**
We illustrate the cross-ancestry genetic correlation between sleep and asthma. The red bars represent genetic correlations between sleep and asthma within the European population, while the blue bars represent genetic correlations between sleep (European) and asthma (East Asian). Sleep traits whose genetic correlations pass the false discovery (FDR) rate of 5% ($P < 1.81 \times 10^{-2}$) with asthma within the European population were plotted.

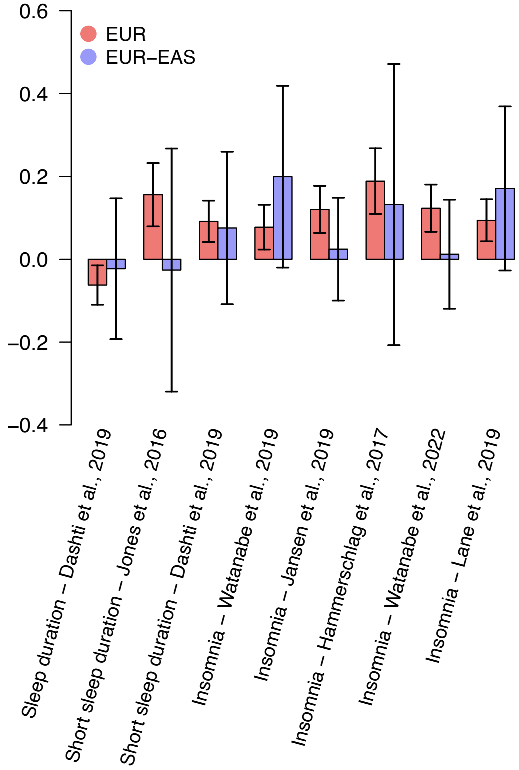


**Fig. S32 Cross-ancestry genetic correlations between sleep and atrial fibrillation.**
We illustrate the cross-ancestry genetic correlation between sleep and atrial fibrillation. The red bars represent genetic correlations between sleep and asthma within the European population, while the blue bars represent genetic correlations between sleep (European) and atrial fibrillation (East Asian). Sleep traits whose genetic correlations pass the false discovery (FDR) rate of 5% ($P < 1.81 \times 10^{-2}$) with atrial fibrillation within the European population were plotted.

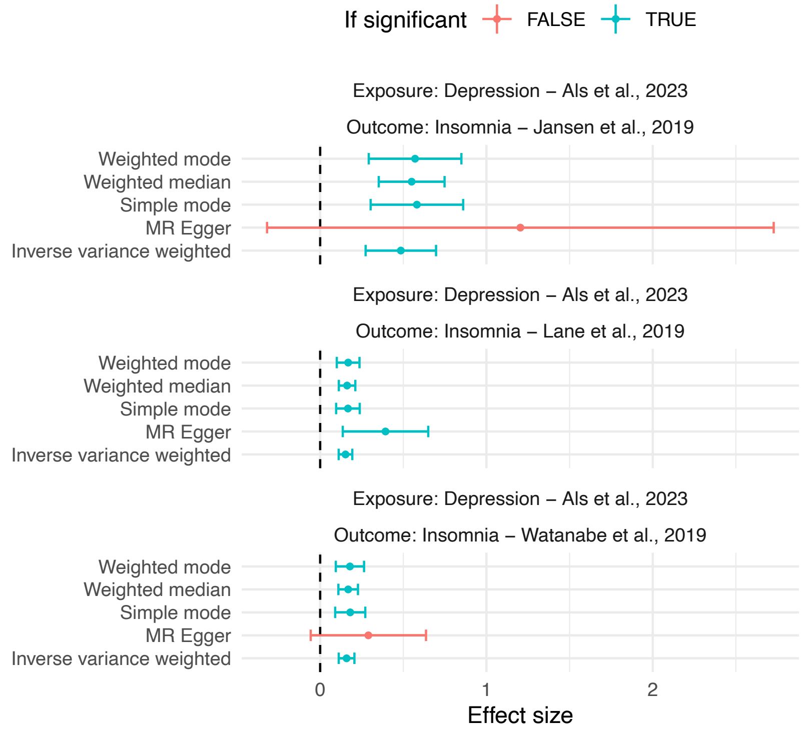


**Fig. S33 Mendelian randomization (MR) results with exposure being depression and outcome being insomnia.**

We illustrate results of MR of five methods with exposure being depression and outcome being insomnia, with different GWAS summary statistics (showed in the title of each panel). Colors represent if the pair is significant with the method specified on the left-hand side.

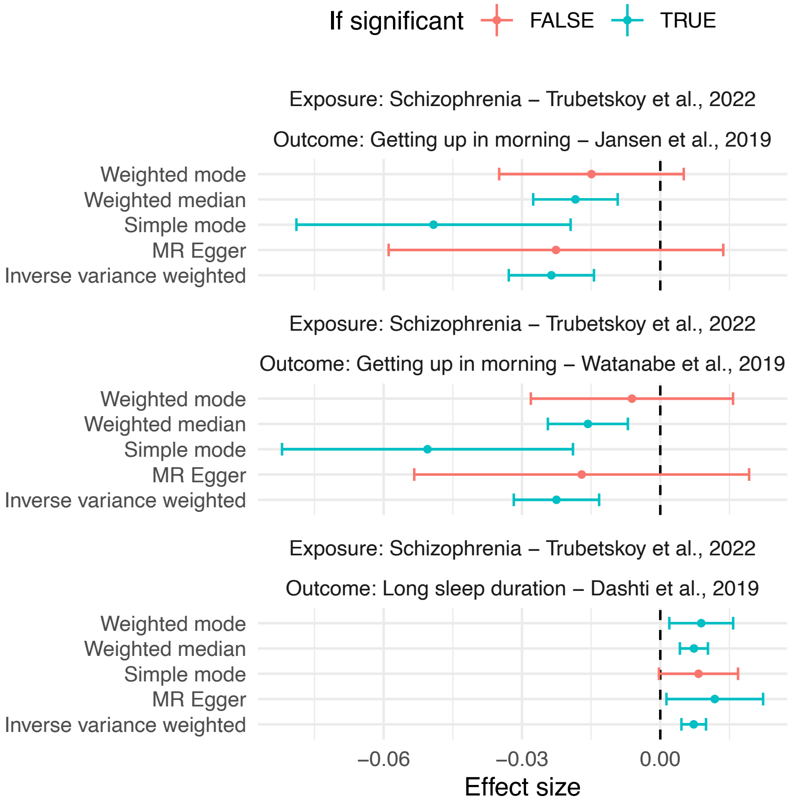


**Fig. S34 Mendelian randomization (MR) results with exposure being schizophrenia and outcome being sleep traits.**

We illustrate results of MR of five methods with exposure being schizophrenia and outcome being (**A**) ease of getting up in the morning; and (**B**) long sleep duration, with different GWAS summary statistics (showed in the title of each panel). Colors represent if the pair is significant with the method specified on the left-hand side.

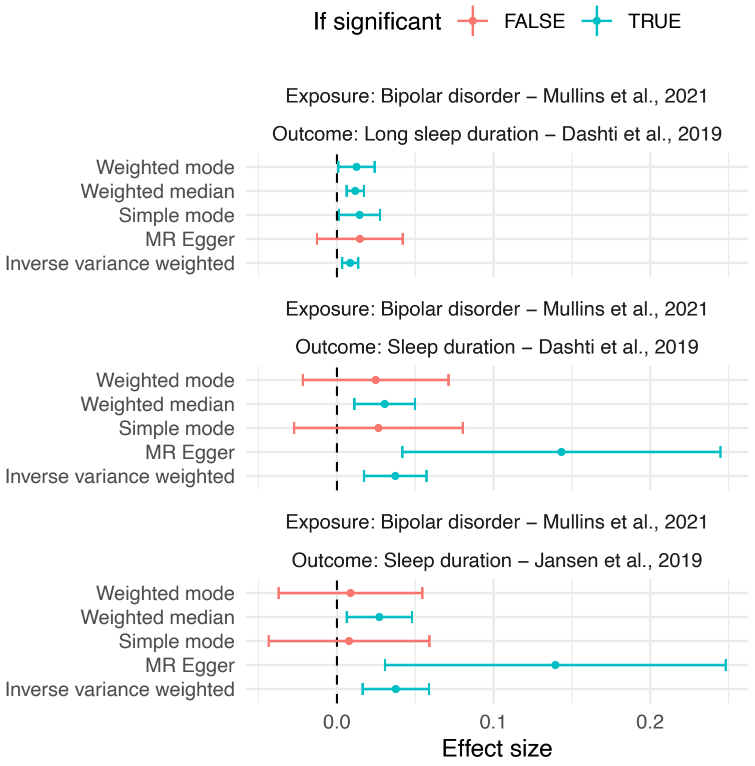


**Fig. S35 Mendelian randomization (MR) results with exposure being bipolar disorder and outcome being sleep duration.**

We illustrate results of MR of five methods with exposure being bipolar disorder and outcome being sleep duration, with different GWAS summary statistics (showed in the title of each panel). Colors represent if the pair is significant with the method specified on the left-hand side.

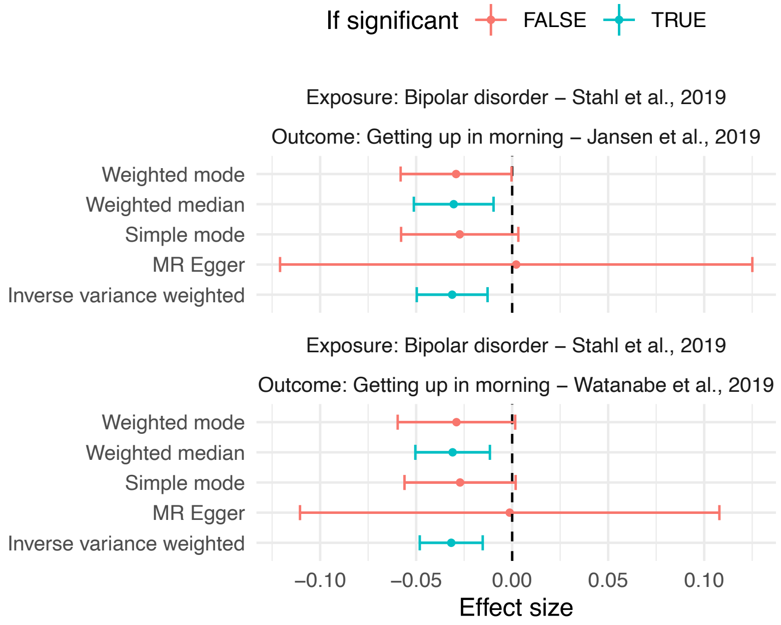


**Fig. S36 Mendelian randomization (MR) results with exposure being bipolar disorder and outcome being ease of getting up in the morning.**

We illustrate results of MR of five methods with exposure being bipolar disorder and outcome being ease of getting up in the morning, with different GWAS summary statistics (showed in the title of each panel). Colors represent if the pair is significant with the method specified on the left-hand side.

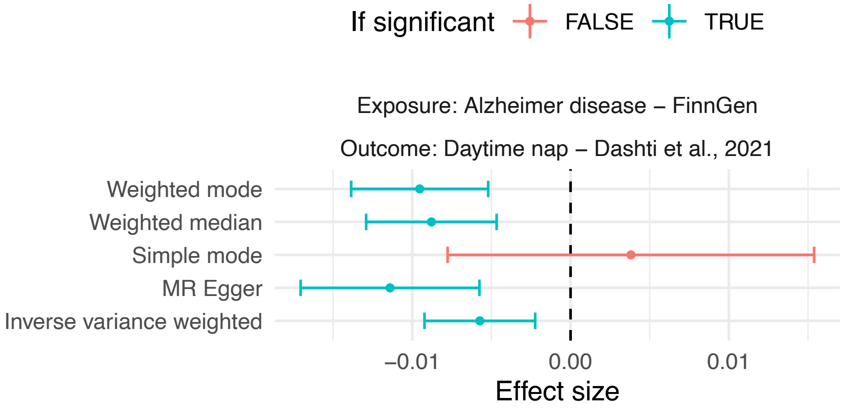


**Fig. S37 Mendelian randomization (MR) results with exposure being Alzheimer's disease and outcome being daytime napping.**

We illustrate results of MR of five methods with exposure being Alzheimer's disease and outcome being daytime napping. Colors represent if the pair is significant with the method specified on the left-hand side.

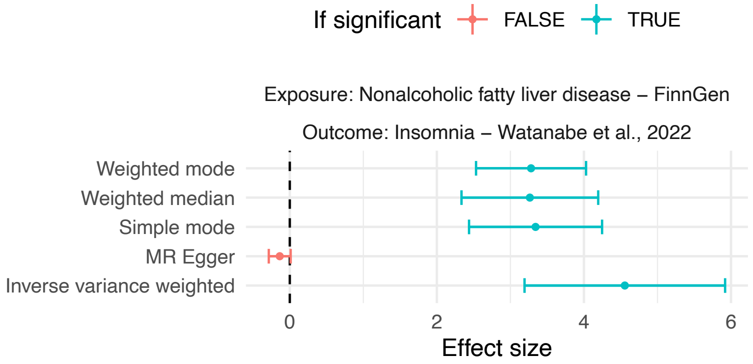


**Fig. S38 Mendelian randomization (MR) results with exposure being nonalcoholic fatty liver disease and outcome being insomnia.**

We illustrate results of MR of five methods with exposure being nonalcoholic fatty liver disease and outcome being insomnia. Colors represent if the pair is significant with the method specified on the left-hand side.

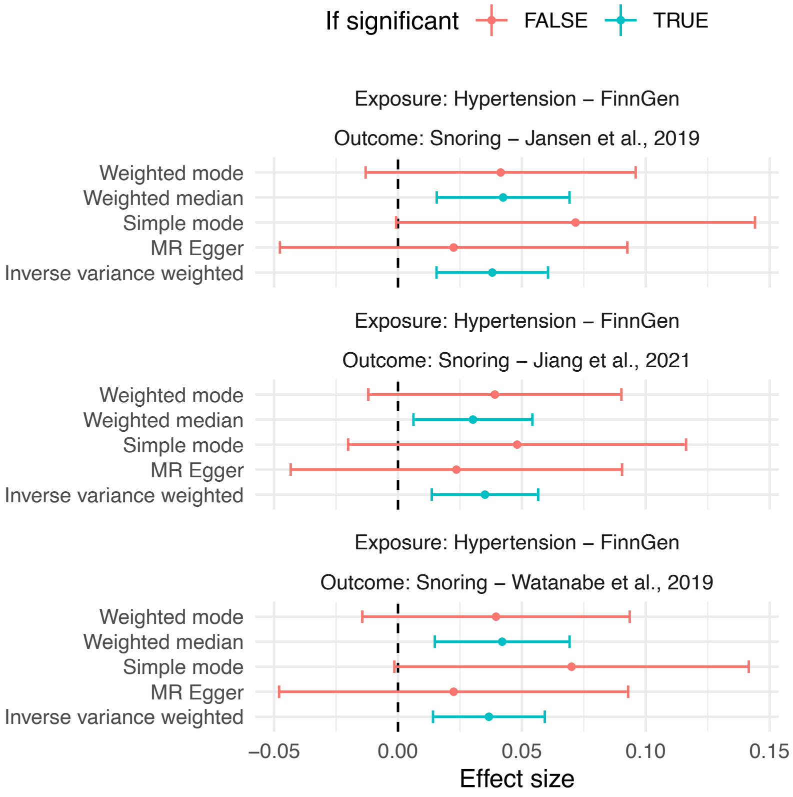


**Fig. S39 Mendelian randomization (MR) results with exposure being hypertension and outcome being snoring.**

We illustrate results of MR of five methods with exposure being hypertension and outcome being snoring, with different GWAS summary statistics (showed in the title of each panel). Colors represent if the pair is significant with the method specified on the left-hand side.

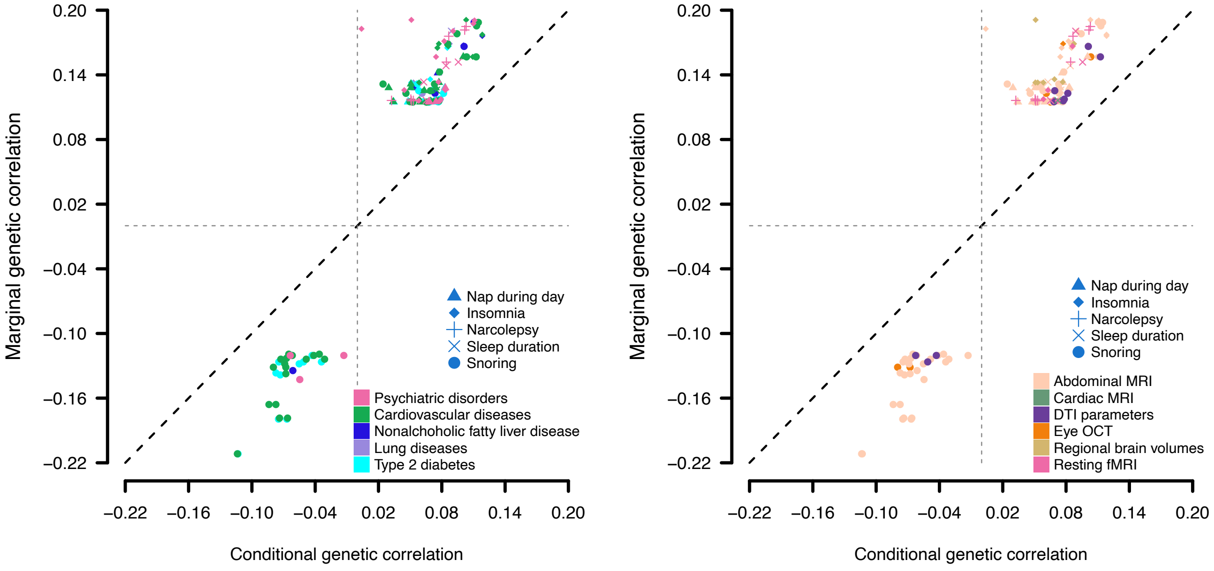


**Fig. S40 Mediation effects of diseases on the associations of imaging and sleep traits.**

We illustrate marginal genetic correlations between imaging and sleep traits (*y* axis) and genetic correlations between imaging and sleep traits mediated by disease (*x* axis). Different sleep traits are represented by different shapes, while each disease category (left panel) and imaging category (right panel) is denoted by a unique color. Only associations with consistent directions of marginal and conditional genetic correlations are plotted.

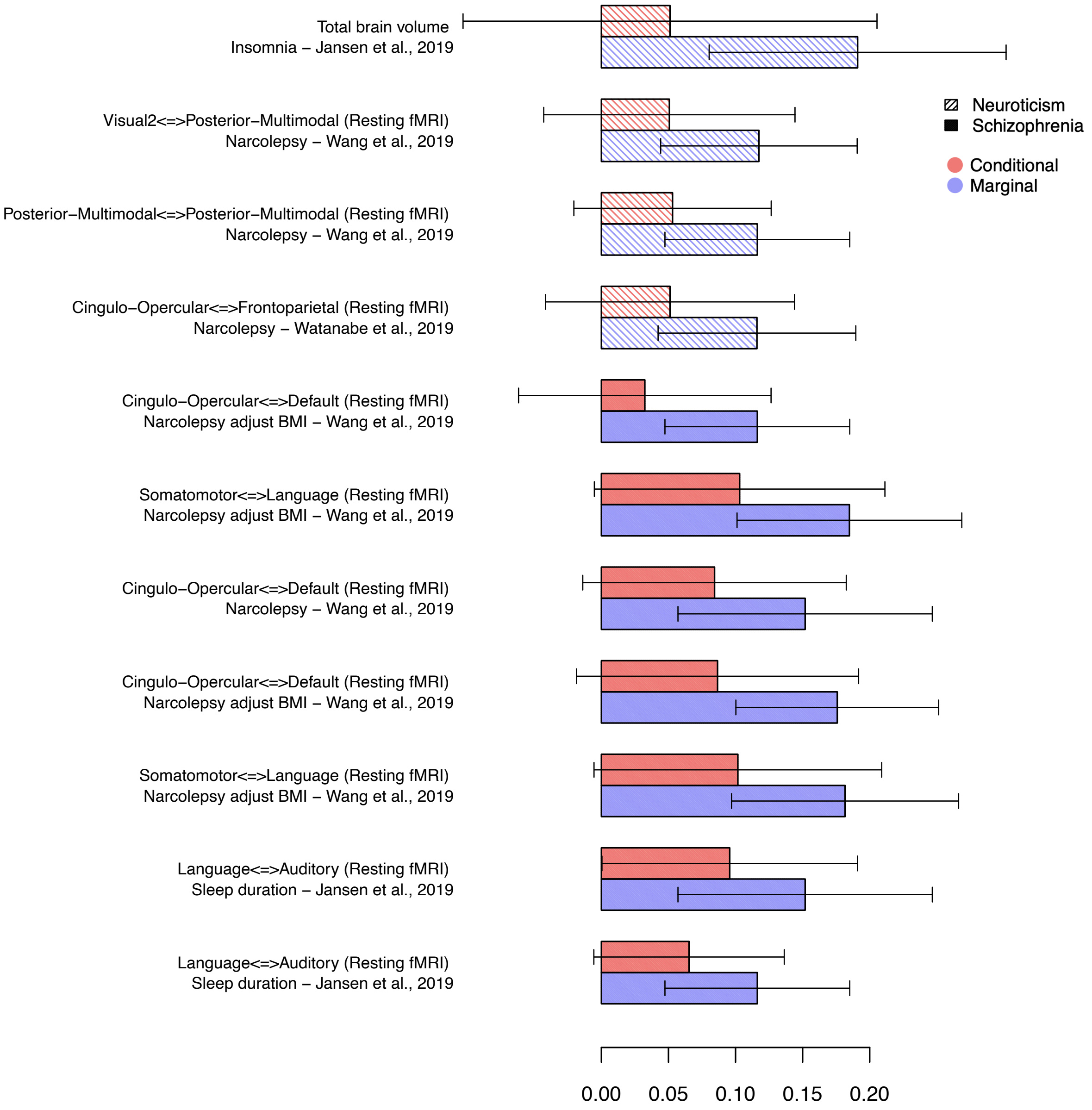


**Fig. S41 Mediation effects of psychiatric disorders on specific associations between brain imaging and sleep traits.**

We illustrate both the marginal genetic correlations between brain imaging and sleep traits, and the genetic correlations between brain imaging and sleep traits mediated by psychiatric disorders. These correlations are distinguished by different colors. The pairs of brain imaging and sleep traits are listed on the left-hand side, while the density of color denotes the different psychiatric disorder.

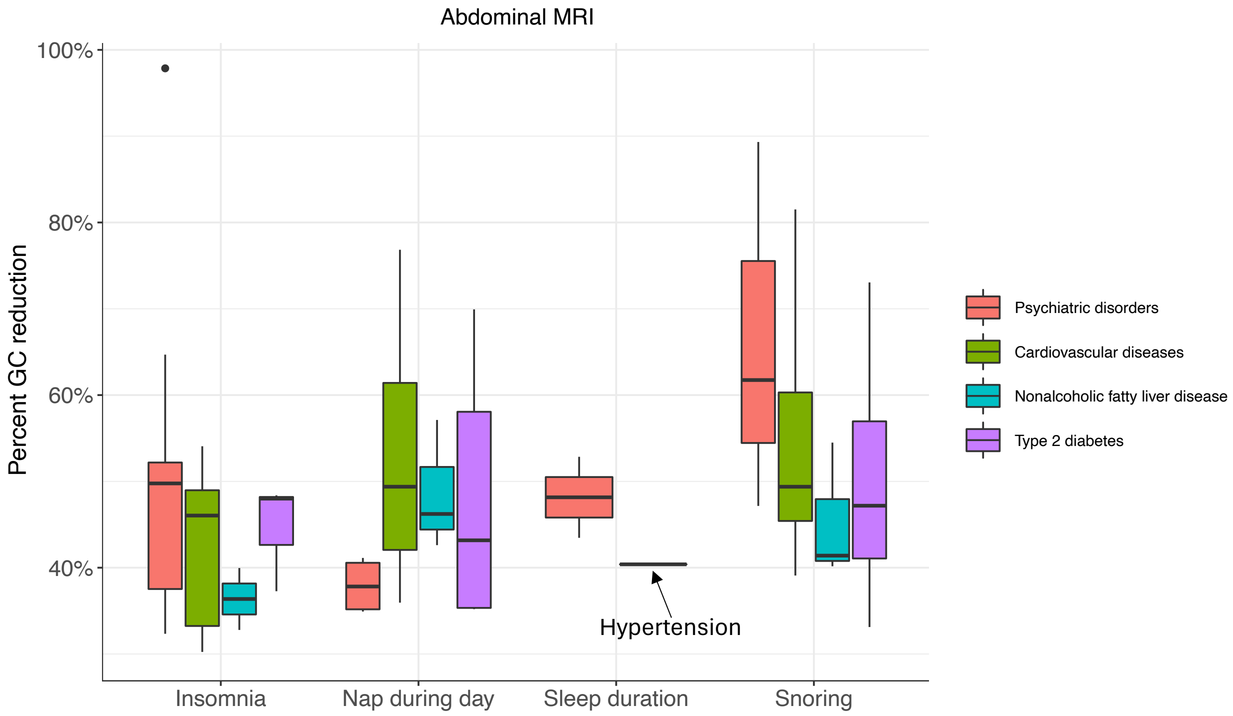


**Fig. S42 Mediation effects of diseases on associations between abdominal MRI and sleep traits.**

We illustrate the percentage reduction in genetic correlation between abdominal MRI and sleep traits, both with and without mediation by diseases. This percentage reduction is calculated as the difference between the marginal genetic correlation and the conditional genetic correlation, divided by the marginal genetic correlation. Each disease category is represented by a distinct color. Only associations with consistent directions of marginal and conditional genetic correlations are plotted.

**Legends for Tables S1 to S13** (All tables can be found in a zip file).

**Table S1. The ID of imaging traits used in phenotypic and genetic association analyses.**

**Table S2. Phenotypic associations between imaging traits and sleep traits.**

**Table S3. Sleep GWAS summary statistics used in the genetic correlation analysis.**

**Table S4. Genetic correlations between imaging traits and sleep traits.**

**Table S5. Independent significant variants and their correlated variants for imaging traits that have previously been identified at P-value < 9e-6 in GWAS of sleep-related traits and disorders listed in the GWAS catalog (version r2022-07-09, www.ebi.ac.uk/gwas/).** See the method section for details on how independent significant variants were defined for each imaging modality.

**Table S6. Summary of the shared genetic effects between multi-organ imaging traits and sleep traits.**

**Table S6. Validating significant CMR associations in independent UKB datasets.**

**Table S7. GWAS summary statistics of multi-organ-related disorders/traits used in the genetic correlation analysis.**

**Table S8. Genetic correlations between sleep and multi-organ-related disorders/traits.**

**Table S9. GWAS summary statistics multi-organ-related disorders/traits used in the cross-population correlation analysis.**

**Table S10. Cross-ancestry genetic correlations between sleep and multi-organ-related diseases/traits.**

**Table S11. GWAS summary statistics of multi-organ-related disorders/traits used in the Mendelian randomization analysis.**

**Table S12. Causal genetic relationships detected by Mendelian randomization.** Significant pairs, determined via inverse variance weighted method after FDR correction ($P < 2.93\times 10^{-3}$), were documented in both Disease_to_sleep_MR_FDR and Sleep_to_disease_MR_FDR sheets, with all five methods listed. Pairs with $P$ values below 0.05 were emphasized in bold.

**Table S13. The mediation effect of disease on the associations between imaging and sleep traits.**
